# Longitudinal associations between weight indices, cognition, and mental health from childhood to early adolescence

**DOI:** 10.1101/2023.12.14.23299828

**Authors:** Zhaolong Adrian Li, Mary Katherine Ray, Yueping Gu, Deanna M. Barch, Tamara Hershey

**Affiliations:** Department of Psychiatry, Washington University School of Medicine in St. Louis, St. Louis, MO 63110, USA; Department of Psychological & Brain Sciences, Washington University in St. Louis, St. Louis, MO 63130, USA

## Abstract

Childhood obesity has been associated with lower cognitive performance and worse mental health in cross-sectional studies. However, it is unclear whether these findings extend longitudinally and in what causal direction. Using data from the Adolescent Brain Cognitive Development (ABCD) Study (maximum analytical n = 6671, 48.3% girls, 42.8% non-White), we examined how body mass index (BMI) at baseline (ages 9-11) relate prospectively to changes in cognition or psychopathology across the 2 years thereafter, and vice versa. Cognitive tests included the National Institutes of Health Toolbox Cognition Battery, Little Man Task of mental rotation, and Rey Auditory Verbal Learning Test. Psychopathology was assessed using caregiver-reported Child Behavior Checklist. Linear mixed models adjusted for sociodemographic and developmental covariates indicated that lower baseline performance on most cognitive tests was associated with greater longitudinal BMI gain (eg, 1 point lower than median on Picture Vocabulary corresponded to 0.012 kg/m^2^ [1.6%; 95% CI, 0.008 to 0.016 kg/m^2^] more annual BMI gain, *P*_FDR_ < .001), whereas baseline BMI was unrelated to longitudinal changes in cognition (*P*_FDR_ ≥ .12; including after considering practice effects). Greater broad-spectrum psychopathology at baseline was associated with increased BMI gain (eg, each endorsement of externalizing problems than none corresponded to 0.015 kg/m^2^ [2.2%; 95% CI, 0.009 to 0.021 kg/m^2^] more annual BMI gain, *P*_FDR_ < .001) and, reciprocally, greater baseline BMI was linked specifically to more longitudinal withdrawn/depressed and depression problems (0.010 [22%; 95% CI, 0.004 to 0.016] and 0.011 [15%; 95% CI, 0.004 to 0.017] more problems annually per 1 kg/m^2^ above median BMI, *P*_FDR_ = .003 and .008). The associations did not differ in boys vs. girls (*P*_FDR_ ≥ .40), and remained stable with waist circumference as the weight index and in subgroups of participants without weight-altering medications or common baseline psychiatric diagnoses. Our longitudinal findings expand previous cross-sectional works and highlight the importance of cognitive and mental health to children’s weight development and links between weight and depression.

## Introduction

High child and adolescent obesity rates (eg, 19.7% in the US) are problematic given links between early-life obesity and long-term health issues^1^. While evidence suggests cross-sectional associations between obesity, lower cognitive functioning, and worse mental health in youth^2^, it remains unclear whether these findings extend longitudinally and in what direction. Leveraging data from the large Adolescent Brain Cognitive Development (ABCD) Study (release 5.0)^3,4^, we examined how weight indices at ages 9-11 relate prospectively to changes in cognition and psychopathology across the 2 years thereafter, and vice versa.

## Methods

Baseline (June 2016 to October 2018) and 1- and 2-year follow-up ABCD Study data collected pre-COVID-19 (March 13, 2020) were included for participants without severe medical conditions (**eMethods**). Weight indices (body mass index [BMI]; waist circumference [WC]) and psychopathology (Child Behavior Checklist) were assessed annually^3^; cognition (National Institutes of Health Toolbox Cognition Battery; Little Man Task of mental rotation; Rey Auditory Verbal Learning Test) was assessed at baseline and 2-year^4^ (**Table**). Caregivers and children provided written informed consent or assent to procedures approved by site institutional review boards. We followed the STROBE reporting guidelines.

We used [age] × [baseline predictor] interactions in linear mixed models to estimate associations between baseline BMI or WC and changes in cognition or psychopathology across timepoints, and vice versa. Models also included lower-order main effects, sociodemographic and developmental covariates, random intercepts of participants within families within sites, and participant-level random slopes (except for cognition-as-outcome models limited by two timepoints) (**Figure** caption). Sensitivity analyses explored sex interactions and whether associations remained in children not using weight-related medications or without common baseline psychiatric diagnoses.

**Figure 1.**
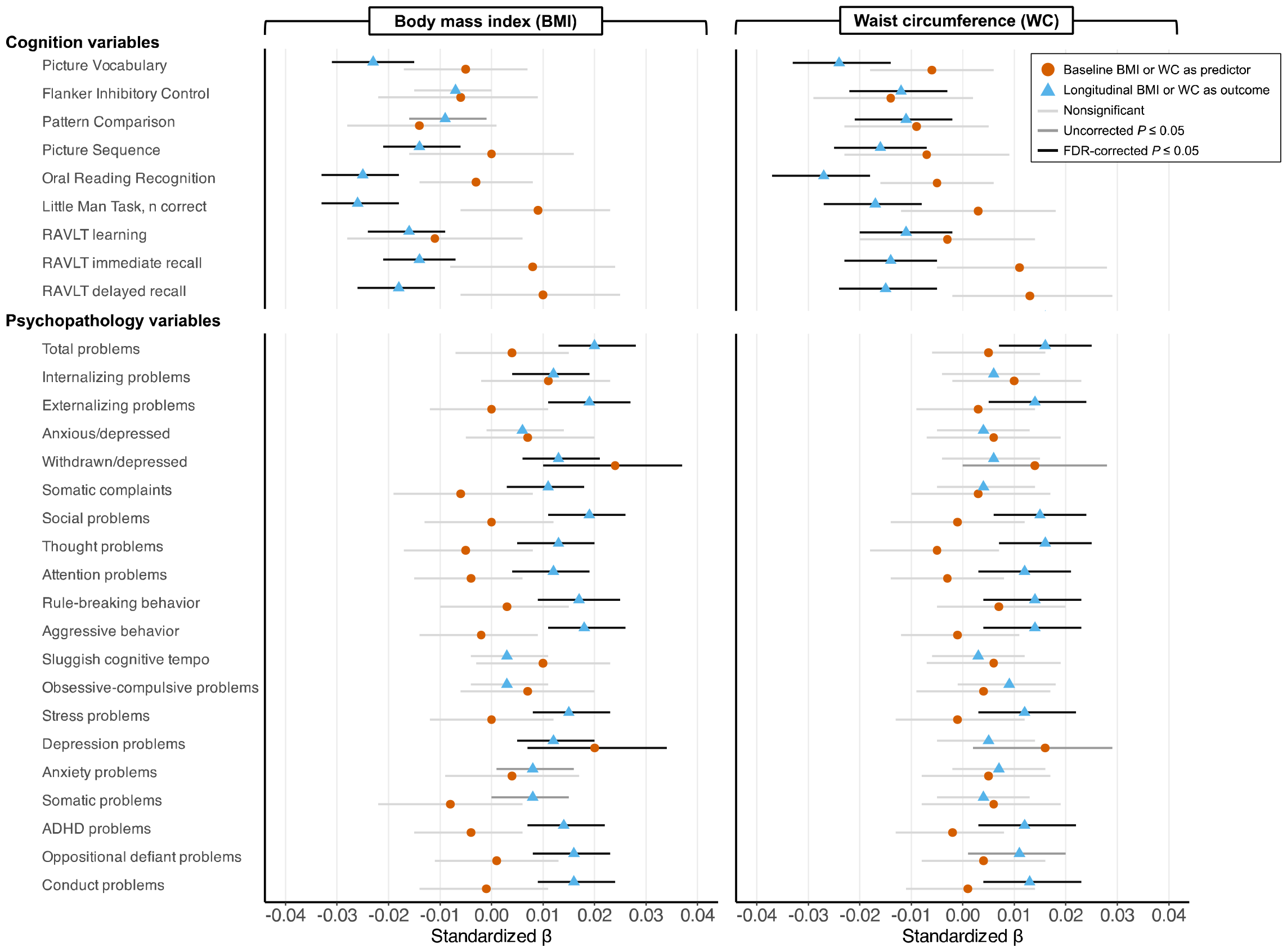
Associations between weight indices, cognition, and psychopathology as children enter adolescence. Standardized β coefficients with 95% profile likelihood confidence intervals were longitudinal [age (centered at median baseline age of 10 y)] × [baseline BMI or WC] interactions (orange circles) or [age] × [baseline cognition or psychopathology] interactions (blue triangles), estimated in linear mixed models. Lower-order main effects were automatically added. Detailed statistics are shown in **eTables 1** (BMI) and **6** (WC), and unstandardized coefficients along with main effects of age are shown in **eTables 2** (BMI) and **7** (WC). Models used the BOBYQA optimizer with 100,000 maximum iterations. Fixed-effect covariates included child sex, area deprivation index national percentile, income-to-needs ratio, pubertal development scale total score, and familial history of mental illness (depression, mania, psychosis, suicide attempt, antisocial behavior) and drug or alcohol use problems. Random intercepts were participants nested within families within study sites. To capture heterogeneity in longitudinal changes, participant-level random slopes were included, except for models where cognition was the outcome and assessed only twice. Multiple comparison correction was performed within domains (cognition or psychopathology) and weight indices (BMI or WC) using false discovery rate (FDR) at two-tailed *P* ≤-.05. ADHD indicates attention-deficit/hyperactivity disorder; RAVLT, Rey Auditory Verbal Learning Test.

**Table 1.** Participant characteristics.

**Table. Participant characteristics**
| Variable<sup>b</sup> | Present study (n = 6671)<sup>a</sup> |  |  |
| --- | --- | --- | --- |
|  | Baseline, mean (SD) | 1-y follow-up, mean (SD) | 2-y follow-up, mean (SD) |
| Age, mean (SD) [range], y | 10.0 (0.6)<br>[8.9 to 11.1] | 11.0 (0.6)<br>[9.8 to 12.4] | 11.9 (0.6)<br>[10.6 to 13.7] |
| Sex |  |  |  |
| Female, No. (%) | 3221 (48.3) | NA | NA |
| Male, No. (%) | 3450 (51.7) | NA | NA |
| Race and ethnicity |  |  |  |
| Asian, No. (%) | 156 (2.3) | NA | NA |
| Black, No. (%) | 778 (11.7) | NA | NA |
| Hispanic, No. (%) | 1269 (19.0) | NA | NA |
| White, No. (%) | 3816 (57.2) | NA | NA |
| Other, No. (%)<sup>c</sup> | 652 (9.8) | NA | NA |
| Area deprivation index national percentile<sup>d</sup> | 38.3 (25.9) | NA | NA |
| Income-to-needs ratio<sup>e</sup> | 3.8 (2.4) | NA | NA |
| Pubertal development scale total score<sup>f</sup> | 7.9 (2.3) | 9.0 (3.0) | 10.6 (3.5) |
| BMI, kg/m<sup>2</sup> | 18.5 (3.7) | 19.5 (4.5) | 20.4 (4.7) |
| WC, in | 26.2 (4.0) | 27.5 (4.4) | 28.5 (4.6) |
| NIHTB Picture Vocabulary score | 85.2 (7.9) | NA | 89.2 (8.4) |
| NIHTB Flanker Inhibitory Control score | 94.8 (8.5) | NA | 100.4 (7.2) |
| NIHTB Pattern Comparison score | 88.7 (14.3) | NA | 103.7 (14.9) |
| NIHTB Picture Sequence score | 103.6 (12.1) | NA | 109.7 (12.0) |
| NIHTB Oral Reading Recognition score | 91.3 (6.7) | NA | 94.9 (6.5) |
| Little Man Task, n correct | 19.1 (5.5) | NA | 23.8 (5.8) |
| RAVLT learning, n correct | 11.5 (2.4) | NA | 11.3 (2.3) |
| RAVLT immediate recall, n correct | 9.9 (3.0) | NA | 9.9 (2.6) |
| RAVLT delayed recall, n correct | 9.4 (3.2) | NA | 9.3 (2.8) |
| CBCL total problems | 16.9 (16.5) | 16.3 (15.9) | 15.5 (15.9) |
| CBCL internalizing behavior | 4.8 (5.2) | 4.8 (5.2) | 4.7 (5.3) |
| CBCL externalizing behavior | 4.1 (5.4) | 3.8 (5.1) | 3.7 (5.2) |
| CBCL anxious/depressed | 2.4 (2.9) | 2.4 (2.9) | 2.2 (2.8) |
| CBCL withdrawn/depressed | 0.9 (1.6) | 1.0 (1.7) | 1.1 (1.8) |
| CBCL somatic complaints | 1.5 (1.9) | 1.4 (1.9) | 1.4 (1.9) |
| CBCL social problems | 1.5 (2.1) | 1.4 (2.0) | 1.2 (2.0) |
| CBCL thought problems | 1.5 (2.0) | 1.5 (2.0) | 1.3 (2.0) |
| CBCL attention problems | 2.8 (3.3) | 2.7 (3.3) | 2.6 (3.2) |
| CBCL rule-breaking behavior | 1.1 (1.7) | 1.0 (1.6) | 1.0 (1.7) |
| CBCL aggressive behavior | 3.0 (4.0) | 2.8 (3.8) | 2.7 (3.8) |
| CBCL sluggish cognitive tempo | 0.5 (0.9) | 0.5 (0.9) | 0.5 (0.9) |
| CBCL obsessive-compulsive problems | 1.3 (1.7) | 1.3 (1.7) | 1.2 (1.7) |
| CBCL stress problems | 2.7 (3.1) | 2.7 (3.1) | 2.6 (3.1) |
| CBCL depression problems | 1.2 (1.9) | 1.3 (2.0) | 1.4 (2.1) |
| CBCL anxiety problems | 2.0 (2.3) | 1.9 (2.3) | 1.7 (2.2) |
| CBCL somatic problems | 1.1 (1.5) | 1.0 (1.4) | 1.0 (1.4) |
| CBCL ADHD problems | 2.5 (2.9) | 2.3 (2.8) | 2.2 (2.7) |
| CBCL oppositional defiant problems | 1.7 (2.0) | 1.6 (1.9) | 1.5 (1.9) |
| CBCL conduct problems | 1.1 (2.1) | 1.0 (2.0) | 1.0 (2.1) |

Abbreviations: ADHD, attention-deficit/hyperactivity disorder; BMI, body mass index; CBCL, Child Behavior Checklist; NA, not applicable; NIHTB, National Institutes of Health Toolbox; RAVLT, Rey Auditory Verbal Learning Test; WC, waist circumference.
<sup>a</sup> For some variables, usable data may come from fewer participants due to missing data and/or excluded outliers (weight indices and cognition values that were 4SD or more away from the mean were removed). See **eTables 1, 3-6, 8-10** for exact numbers of participants included in analytical models.
<sup>b</sup> Sociodemographic variables were assessed at baseline only. Cognition was assessed at baseline and 2-y follow-up using uncorrected scores that evaluated domains of verbal and reading ability (Picture Vocabulary, Oral Reading Recognition), executive control and attention (Flanker Inhibitory Control), processing speed (Pattern Comparison), learning and episodic memory (Picture Sequence, RAVLT), and visuospatial processing (Little Man Task)<sup>4</sup>. Higher scores reflect better performance. Annual psychopathology assessments using caregiver-reported CBCL raw scores included 11 empirical syndrome subscales (total problems [summarizing internalizing, externalizing, social, thought, and attention problems], internalizing [summarizing anxious/depressed, withdrawn/depressed, and somatic complaints], externalizing [summarizing rule-breaking and aggressive behavior], and individual constituent subscales), 3 other subscales (sluggish cognitive tempo, obsessive-compulsive problems, and stress problems), and 6 subscales deemed cross-culturally consistent with *Diagnostic and Statistical Manual of Mental Disorders, Fifth Edition* categories (depression, anxiety, somatic, ADHD, oppositional defiant, and conduct problems)<sup>3</sup>. Higher scores reflect greater endorsement.
<sup>c</sup> Includes caregiver-reported American Indian or Alaska Native, Native Hawaiian or other Pacific Islander, multiple races and/or ethnicities, and unknown race or ethnicity.
<sup>d</sup> Based on American Community Survey (2011-2015) estimates at census block level. Higher percentiles reflect greater neighborhood socioeconomic disadvantage.
<sup>e</sup> Calculated as the median level of household income bands divided by the 2017 US federal poverty guidelines given household size.
<sup>f</sup> Based on caregiver-reported pubertal status ratings that previously showed high correlation with Tanner stages<sup>3</sup>. Higher values reflect more advanced pubertal stage.

## Results

Participant (n = 6671) characteristics are shown in **Table**. Baseline BMI was not associated with longitudinal changes in cognition (**Figure**; **eTable 1**), and this finding was not confounded by practice effects from repeated cognitive testing (**eMethods**). Conversely, lower baseline cognition was overall associated with greater longitudinal BMI gain. Unstandardized estimates indicated, eg, that children scoring 1 point lower on Picture Vocabulary at baseline had 0.012 kg/m^2^ (1.6%) more annual BMI gain than those scoring at the median (**eTable 2**).

Higher baseline BMI was associated with more longitudinal withdrawn/depressed and depression problems (**Figure**; **eTable 1**), with each 1 kg/m^2^ increase corresponding to 0.010 (22%) and 0.011 (15%) more problems annually beyond changes at median BMI (**eTable 2**). On the other hand, greater baseline psychopathology was broadly associated with greater BMI gain. For instance, each baseline endorsement of externalizing and social problems corresponded to 0.015 kg/m^2^ (2.2%) and 0.038 kg/m^2^ (5.4%) more annual BMI increases compared to no endorsement.

No interaction with sex was found in any analysis (**eTable 3**). Results remained consistent in subgroups of participants without weight-altering medications (**eTable 4**) or common baseline psychiatric diagnoses (**eTable 5**). Findings with WC were similar to those with BMI, except baseline internalizing spectrum did not predict longitudinal changes in WC (**eTables 6**-**10**).

## Supporting information

eMethods; eTables 1-10; eFigures 1-2

## Data Availability

Data in this study were from the ABCD Release 5.0 (September 26, 2023; https://doi.org/10.15154/8873-zj65). The ABCD Study repository grows and may be modified as new data are collected and processed.

## Discussion

Lower cognitive performance and greater broad-spectrum psychopathology were associated with increased weight gain as children entered adolescence. These factors have been linked to brain reward network dysfunction, impaired reward learning, and caregiver-child conflicts that potentially hamper adherence to healthy diet and lifestyle^2,5,6^. Reciprocally, higher weight was associated with more depression over time. Obesity may aggravate psychological stress via body dissatisfaction and weight-related discrimination, and also trigger inflammation-mediated hypothalamic-pituitary-adrenal axis dysregulation, both of which contribute to depression^2,6^. Together, our longitudinal results clarify previous cross-sectional findings and highlight the importance of cognitive and mental health to children’s weight development and links between weight and depression.

Limitations include the limited longitudinal timeframe, inadequate executive function assessments, and lack of body composition measures. Future ABCD Study data and focused clinical cohorts can further delineate adolescent development, exploring late-emerging effects^5^, nonlinear trends, and biopsychosocial mediators.

## Author Contributions

Mr. Li had full access to all of the data in the study and takes responsibility for the integrity of the data and the accuracy of the data analysis.

*Concept and design*: Li, Ray, Hershey.

*Acquisition, analysis, or interpretation of data*: All authors.

*Drafting of the manuscript*: Li.

*Critical revision of the manuscript for important intellectual content*: All authors.

*Statistical analysis*: Li, Gu. *Obtained funding*: Hershey. *Supervision*: Hershey.

## Disclaimer

The content is solely the responsibility of the authors and does not necessarily represent the official views of the National Institutes of Health or other funders.

## Additional Contributions

The authors thank Amjad Samara, MD (Department of Neurology, Washington University School of Medicine in St. Louis) for discussions about statistical analysis, for which financial compensation was not received.

