## Supplementary material for "Longitudinal associations between weight indices, cognition, and mental health from childhood to early adolescence": eMethods; eTables 1-10; eFigures 1-2

**eMethods.** Participant selection; Practice effects in cognitive scores (pp. 2-4)

**eTable 1.** Longitudinal associations between BMI, cognition, and psychopathology from late childhood through early adolescence (standardized estimates) (p. 5)

**eTable 2.** Longitudinal associations between BMI, cognition, and psychopathology from late childhood through early adolescence (unstandardized estimates) (p. 6)

**eFigure 1.** Johnson-Neyman interaction plots for significant longitudinal associations between BMI and cognition from late childhood through early adolescence (unstandardized estimates) (p. 7)

**eFigure 2.** Johnson-Neyman interaction plots for significant longitudinal associations between BMI and psychopathology from late childhood through early adolescence (unstandardized estimates) (pp. 8-9)

**eTable 3.** Sex interactions with the longitudinal associations between BMI, cognition, and psychopathology from late childhood through early adolescence (p. 10)

**eTable 4.** Longitudinal associations between BMI, cognition, and psychopathology in children not using weight-related medications (p. 11)

**eTable 5.** Longitudinal associations between BMI, cognition, and psychopathology in children without caregiver-reported common psychiatric diagnoses at baseline (p. 12)

**eTable 6.** Longitudinal associations between WC, cognition, and psychopathology from late childhood through early adolescence (standardized estimates) (p. 13)

**eTable 7.** Longitudinal associations between WC, cognition, and psychopathology from late childhood through early adolescence (unstandardized estimates) (p. 14)

**eTable 8.** Sex interactions with the longitudinal associations between WC, cognition, and psychopathology from late childhood through early adolescence (p. 15)

**eTable 9.** Longitudinal associations between WC, cognition, and psychopathology in children not using weight-related medications (p. 16)

**eTable 10.** Longitudinal associations between WC, cognition, and psychopathology in children without caregiver-reported common psychiatric diagnoses at baseline (p. 17)

**References.** (p. 18)

**eMethods.** Participant selection; Practice effects in cognitive scores

**Participant selection.** Details on the Adolescent Brain Cognitive Development (ABCD) Study recruitment have been published elsewhere^1^. The ABCD Study was conducted at 21 US sites and children were enrolled using a school-based recruitment system. The recruitment catchment of these sites was demographically representative of the US national and encompassed over 20% of US children aged 9-11 years. Children were included in the ABCD Study if they were 9-11 years old at baseline (June 2016 to October 2018) and fluent in English. Children who had history of severe neurological or psychiatric disorders, magnetic resonance imaging (MRI) contraindications, or were born prematurely (30 days) were excluded. The baseline ABCD cohort included 11864 children. In the present study, we additionally excluded children who had caregiver-reported history of cerebral palsy, brain tumor, brain aneurysm, brain hemorrhage, brain hematoma, stroke, epilepsy, seizures, traumatic brain injury, lead poisoning, multiple sclerosis, muscular dystrophy, intellectual disability, substance use disorder, schizophrenia, autism spectrum disorder, and other serious neurological or psychiatric conditions (n = 666). We further excluded children with diabetes (n = 81), eating disorders including anorexia nervosa, bulimia nervosa, and binge eating at present, in the past, or in remission (n = 241), or with casts/prostheses (n = 82). Lastly, given evidence of COVID-19-related weight gain in children^2^ and disruptions in study administration, we included only participants whose baseline, 1-y, and 2-y data were all collected in-person before March 13, 2020 (date of US declaration of national emergency). The final maximum analytical sample included 6671 children.

**Practice effects in cognitive scores.** Repeated participation in cognitive tests may yield experience-driven performance improvement that confound with changes due to brain and cognitive development. Although the ABCD Study cognitive assessment frequency (every other year) and tasks were designed to minimize practice effects^3^, Anokhin et al. had reported significant practice effects seen at 2-y follow-up relative to baseline^4^. In the present study, we replicated their analyses by comparing cognitive performance in age-matched pairs where one participant came from baseline (first assessment) and the other from 2-y follow-up (second assessment). A total of 1697 children were in the age overlap of 10.6 y (127 mo) to 11.1 y (133 mo) across the two timepoints (**eMethods Figure** panel **A**), of which 612 pairs had matching age, sex, race and ethnicity, area deprivation index national percentile, income-to-needs ratio, baseline age- and sex-adjusted body mass index (BMI) and waist circumference (WC) z-scores, and baseline pubertal development scale total score established using 1-to-1 Mahalanobis distance matching (**eMethods Figure** panel **B**).

We estimated practice effects in cognitive scores by subtracting the baseline data from the 2-y follow-up data in each matched pair, finding very similar values to the estimates reported by Anokhin et al. (**eMethods Table 1**)^4^. These estimates were potentially generalizable to our entire study sample (n = 6671), because the 2-y follow-up participants (who demonstrated practice effects) had similar characteristics compared to all included participants except in terms of income-to-needs ratio. Fisher’s exact test or Wilcoxon rank sum test showed: sex (*P* = .98), race and ethnicity (*P* = .27), area deprivation index national percentile (*P* = .67), income-to-needs ratio (*P* = .001), baseline BMI z-score (*P* = .38), baseline WC z-score (*P* = .96).

**eMethods Figure.**
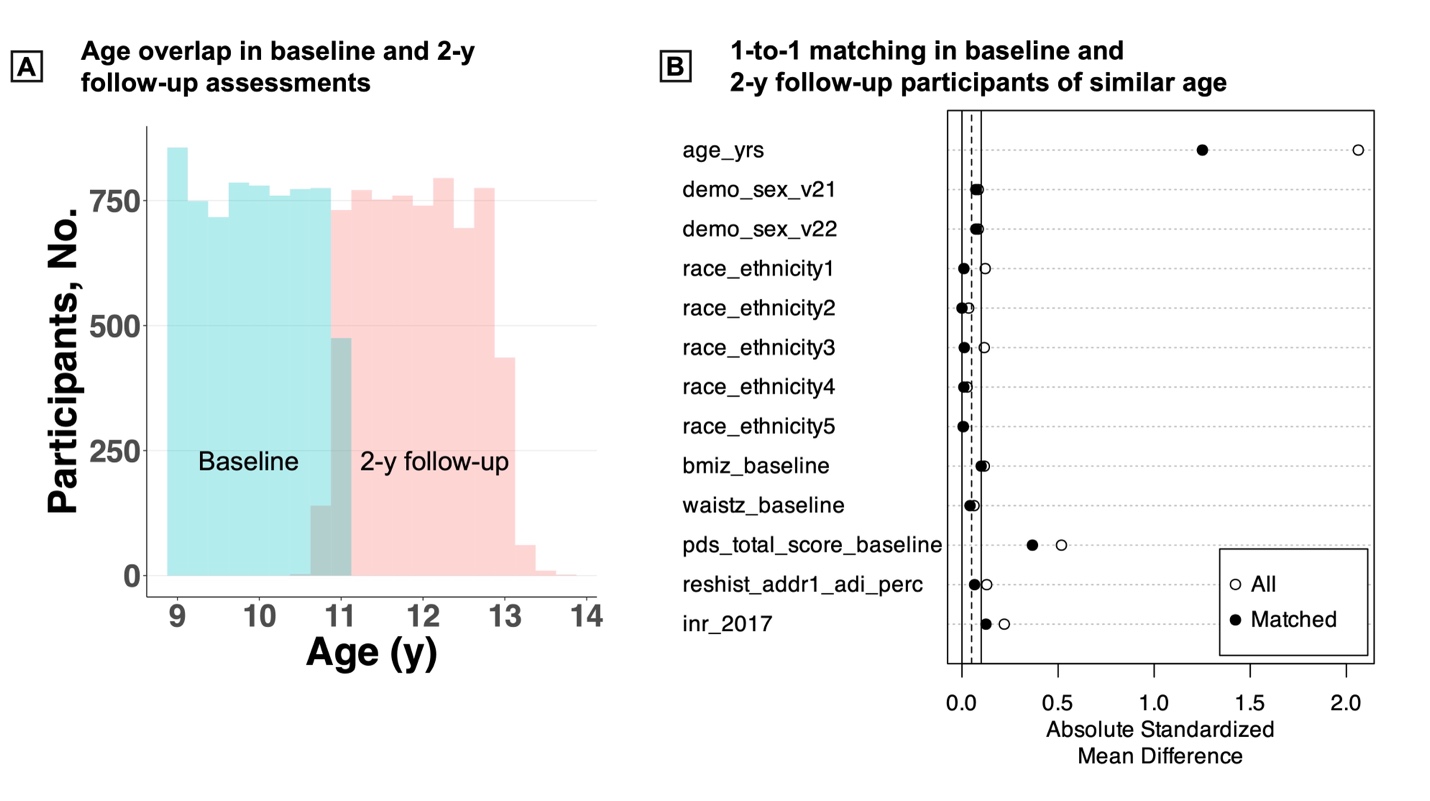

*Caption.* Note the age difference shown in panel **B** among matched pairs was small (0.11 y, or 1.3 mo) even though there was a noticeable absolute standardized mean difference.

**eMethods Table 1.** Estimated practice effects (n = 612)

| **Variable** | **Mean (SD)** | **Median** | **Anokhin et al.**^4^ |
| --- | --- | --- | --- |
| NIHTB Picture Vocabulary score | -0.85 (10.00) | -1 | -0.84 |
| NIHTB Flanker Inhibitory Control score | 1.39 (10.60) | 1 | 1.94 |
| NIHTB Pattern Comparison score | 4.58 (18.88) | 4 | 5.87 |
| NIHTB Picture Sequence score | 2.37 (17.02) | 3 | 2.93 |
| NIHTB Oral Reading Recognition score | 0.14 (8.43) | 0 | -0.43 |
| Little Man Task, n correct | 1.44 (7.97) | 1 | 1.97 |
| RAVLT learning, n correct | -0.66 (3.22) | -1 | -0.75 |
| RAVLT immediate recall, n correct | -0.67 (3.71) | -1 | -0.84 |
| RAVLT delayed recall, n correct | -0.79 (3.92) | -1 | -0.89 |

Importantly, our practice effects were mostly uncorrelated with baseline BMI or WC (**eMethods Table 2**); ie, children gained the same amount of improvement from repeated assessments irrespective of their baseline weight status, consistent with a prior finding^5^. Therefore, any association between baseline weight indices and longitudinal changes in cognition would likely be driven by age-related developmental changes and not practice effects.

**eMethods Table 2.** Pearson correlations between baseline weight and practice effects

| **Estimated practice effects** | **Baseline BMI** | | **Baseline WC** | |
| --- | --- | --- | --- | --- |
|  | ***r*** | ***P* value** | ***r*** | ***P* value** |
| NIHTB Picture Vocabulary score | -0.03 | .41 | 0.01 | .79 |
| NIHTB Flanker Inhibitory Control score | -0.10 | .02 | -0.02 | .55 |
| NIHTB Pattern Comparison score | -0.01 | .77 | 0.00 | .96 |
| NIHTB Picture Sequence score | 0.00 | .99 | 0.01 | .88 |
| NIHTB Oral Reading Recognition score | -0.01 | .76 | 0.03 | .44 |
| Little Man Task, n correct | 0.01 | .87 | 0.04 | .29 |
| RAVLT learning, n correct | -0.02 | .61 | 0.00 | .93 |
| RAVLT immediate recall, n correct | 0.05 | .21 | 0.09 | .03 |
| RAVLT delayed recall, n correct | 0.05 | .21 | 0.07 | .06 |

**eTable 1.** Longitudinal associations between BMI, cognition, and psychopathology from late childhood through early adolescence (standardized estimates)^a^

| **Variable^b^** | **Baseline BMI predicting longitudinal cognition or psychopathology**  (ie, [age] × [baseline BMI] interaction) | | | | **Baseline cognition or psychopathology predicting longitudinal BMI**  (ie, [age] × [baseline cognition or psychopathology] interaction) | | | |
| --- | --- | --- | --- | --- | --- | --- | --- | --- |
|  | **Std. *β* (95% CI)** | ***P* value (raw)** | ***P* value (FDR)** | **n** | **Std. *β* (95% CI)** | ***P* value (raw)** | ***P* value (FDR)** | **n** |
| **Cognition** | | | | | | | | |
| Picture Vocabulary | -0.005 (-0.017 to 0.007) | .42 | .49 | 5198 | -0.023 (-0.031 to 0.015) | < .001 | < .001 | 5212 |
| Flanker Inhibitory Control | -0.006 (-0.022 to 0.009) | .43 | .49 | 5204 | -0.007 (-0.015 to 0.000) | .06 | .12 | 5208 |
| Pattern Comparison | -0.014 (-0.028 to 0.001) | .06 | .12 | 5203 | -0.009 (-0.016 to -0.001) | .02 | .05 | 5201 |
| Picture Sequence | 0.000 (-0.016 to 0.016) | .98 | .98 | 5203 | -0.014 (-0.021 to -0.005) | < .001 | .001 | 5209 |
| Oral Reading Recognition | -0.003 (-0.014 to 0.008) | .55 | .59 | 5197 | -0.025 (-0.033 to -0.018) | < .001 | < .001 | 5204 |
| Little Man Task, n correct | 0.009 (-0.006 to 0.023) | .24 | .33 | 5196 | -0.026 (-0.033 to -0.018) | < .001 | < .001 | 5082 |
| RAVLT learning | -0.011 (-0.028 to 0.006) | .20 | .33 | 5196 | -0.016 (-0.024 to -0.009) | < .001 | < .001 | 5193 |
| RAVLT immediate recall | 0.008 (-0.008 to 0.024) | .35 | .44 | 5193 | -0.014 (-0.021 to -0.007) | < .001 | .001 | 5183 |
| RAVLT delayed recall | 0.010 (-0.006 to 0.025) | .23 | .33 | 5192 | -0.018 (-0.026 to -0.011) | < .001 | < .001 | 5170 |
| **Psychopathology** | | | | | | | | |
| Total problems | 0.004 (-0.007 to 0.015) | .50 | .63 | 5242 | 0.020 (0.013 to 0.028) | < .001 | < .001 | 5269 |
| Internalizing problems | 0.011 (-0.002 to 0.023) | .10 | .18 | 5242 | 0.012 (0.004 to 0.019) | .002 | .006 | 5269 |
| Externalizing problems | 0.000 (-0.012 to 0.011) | .94 | .98 | 5242 | 0.019 (0.011 to 0.027) | < .001 | < .001 | 5269 |
| Anxious/depressed | 0.007 (-0.005 to 0.020) | .25 | .41 | 5242 | 0.006 (-0.001 to 0.014) | .10 | .18 | 5269 |
| Withdrawn/depressed | 0.024 (0.010 to 0.037) | < .001 | .003 | 5242 | 0.013 (0.006 to 0.021) | < .001 | .002 | 5269 |
| Somatic complaints | -0.006 (-0.019 to 0.008) | .40 | .56 | 5242 | 0.011 (0.003 to 0.018) | .004 | .01 | 5269 |
| Social problems | 0.000 (-0.013 to 0.012) | .96 | .98 | 5242 | 0.019 (0.011 to 0.026) | < .001 | < .001 | 5269 |
| Thought problems | -0.005 (-0.017 to 0.008) | .48 | .61 | 5242 | 0.013 (0.005 to 0.020) | < .001 | .003 | 5269 |
| Attention problems | -0.004 (-0.015 to 0.006) | .41 | .56 | 5242 | 0.012 (0.004 to 0.019) | .002 | .006 | 5269 |
| Rule-breaking behavior | 0.003 (-0.010 to 0.015) | .65 | .77 | 5242 | 0.017 (0.009 to 0.025) | < .001 | < .001 | 5269 |
| Aggressive behavior | -0.002 (-0.014 to 0.009) | .68 | .78 | 5242 | 0.018 (0.011 to 0.026) | < .001 | < .001 | 5269 |
| Sluggish cognitive tempo | 0.010 (-0.003 to 0.023) | .14 | .26 | 5242 | 0.003 (-0.004 to 0.011) | .39 | .56 | 5269 |
| Obsessive-compulsive problems | 0.007 (-0.006 to 0.020) | .31 | .50 | 5242 | 0.003 (-0.004 to 0.011) | .38 | .56 | 5269 |
| Stress problems | 0.000 (-0.012 to 0.012) | .98 | .98 | 5242 | 0.015 (0.008 to 0.023) | < .001 | < .001 | 5269 |
| Depression problems | 0.020 (0.007 to 0.034) | .003 | .008 | 5242 | 0.012 (0.005 to 0.020) | .001 | .004 | 5269 |
| Anxiety problems | 0.004 (-0.009 to 0.017) | .53 | .64 | 5242 | 0.008 (0.001 to 0.016) | .03 | .06 | 5269 |
| Somatic problems | -0.008 (-0.022 to 0.006) | .23 | .40 | 5242 | 0.008 (0.000 to 0.015) | .04 | .08 | 5269 |
| ADHD problems | -0.004 (-0.015 to 0.006) | .42 | .56 | 5242 | 0.014 (0.007 to 0.022) | < .001 | .001 | 5269 |
| Oppositional defiant problems | 0.001 (-0.011 to 0.013) | .88 | .95 | 5242 | 0.016 (0.008 to 0.023) | < .001 | < .001 | 5269 |
| Conduct problems | -0.001 (-0.014 to 0.011) | .83 | .93 | 5242 | 0.016 (0.009 to 0.024) | < .001 | < .001 | 5269 |

Abbreviations: ADHD, attention-deficit/hyperactivity disorder; BMI, body mass index; CI, confidence interval; FDR, false discovery rate; RAVLT, Rey Auditory Verbal Learning Test; Std., standardized.

^a^ See **Figure** caption in main text for model specification.

^b^ See notes under **Table** in main text for variable details.

**eTable 2.** Longitudinal associations between BMI, cognition, and psychopathology from late childhood through early adolescence (unstandardized estimates)^a^

| **Variable^b^** | **Baseline BMI predicting longitudinal cognition or psychopathology** | | | **Baseline cognition or psychopathology predicting longitudinal BMI** | | |
| --- | --- | --- | --- | --- | --- | --- |
|  | **Annual change in cognition or psychopathology at median BMI of 17.48 kg/m^2^ (95% CI)**  (ie, main effect of [age]) | **Additional annual change in cognition or psychopathology per 1 kg/m^2^ increase in BMI (95% CI)**  (ie, interaction of [age] × [baseline BMI]) | **%** | **Annual change in BMI at median cognition or no psychopathology (95% CI)**  (ie, main effect of [age]) | **Additional annual change in BMI per 1 point increase in cognition or psychopathology (95% CI)**  (ie, interaction of [age] × [baseline cognition or psychopathology]) | **%** |
| **Cognition** | | | | | | |
| Picture Vocabulary | 2.235 (2.127 to 2.344)^c^ | -0.009 (-0.032 to 0.013) | -0.4 | 0.768 (0.703 to 0.831)^c^ | -0.012 (-0.016 to -0.008)^c^ | -1.6 |
| Flanker Inhibitory Control | 2.740 (2.599 to 2.881)^c^ | -0.012 (-0.043 to 0.018) | -0.4 | 0.758 (0.692 to 0.822)^c^ | -0.004 (-0.007 to 0.000) | -0.5 |
| Pattern Comparison | 7.040 (6.790 to 7.289)^c^ | -0.051 (-0.105 to 0.003) | -0.7 | 0.758 (0.692 to 0.821)^c^ | -0.003 (-0.005 to 0.000) | -0.4 |
| Picture Sequence | 3.244 (3.037 to 3.452)^c^ | 0.001 (-0.045 to 0.046) | 0.0 | 0.766 (0.701 to 0.830)^c^ | -0.005 (-0.007 to -0.002)^c^ | -0.7 |
| Oral Reading Recognition | 1.818 (1.734 to 1.902)^c^ | -0.005 (-0.023 to 0.012) | -0.3 | 0.774 (0.710 to 0.836)^c^ | -0.016 (-0.021 to -0.011)^c^ | -2.1 |
| Little Man Task, n correct | 2.277 (2.183 to 2.370)^c^ | 0.012 (-0.008 to 0.032) | 0.5 | 0.787 (0.720 to 0.852)^c^ | -0.020 (-0.026 to -0.014)^c^ | -2.5 |
| RAVLT learning | 0.082 (0.040 to 0.126)^c^ | -0.006 (-0.016 to 0.003) | -7.3 | 0.738 (0.675 to 0.800)^c^ | -0.028 (-0.040 to -0.015)^c^ | -3.8 |
| RAVLT immediate recall | 0.166 (0.188 to 0.214)^c^ | 0.005 (-0.005 to 0.015) | 3.0 | 0.753 (0.690 to 0.814)^c^ | -0.020 (-0.031 to -0.009)^c^ | -2.7 |
| RAVLT delayed recall | 0.080 (0.030 to 0.130)^c^ | 0.007 (-0.004 to 0.017) | 8.8 | 0.744 (0.680 to 0.806)^c^ | -0.025 (-0.035 to -0.014)^c^ | -3.4 |
| **Psychopathology** | | | | | | |
| Total problems | -0.827 (-1.083 to -0.562^)c^ | 0.016 (-0.031 to 0.063) | -1.9 | 0.670 (0.598 to 0.741)^c^ | 0.005 (0.003 to 0.007)^c^ | 0.7 |
| Internalizing problems | -0.079 (-0.167 to 0.012) | 0.014 (-0.003 to 0.031) | -17 | 0.713 (0.643 to 0.782)^c^ | 0.010 (0.003 to 0.016)^c^ | 1.4 |
| Externalizing problems | -0.297 (-0.378 to -0.213)^c^ | -0.001 (-0.016 to 0.015) | 0.3 | 0.696 (0.626 to 0.763)^c^ | 0.015 (0.009 to 0.021)^c^ | 2.2 |
| Anxious/depressed | -0.080 (-0.129 to -0.030)^c^ | 0.006 (-0.004 to 0.015) | -7.5 | 0.736 (0.667 to 0.804)^c^ | 0.009 (-0.002 to 0.020) | 1.2 |
| Withdrawn/depressed | 0.046 (0.017 to 0.077)^c^ | 0.010 (0.004 to 0.016)^c^ | 22 | 0.725 (0.658 to 0.789)^c^ | 0.037 (0.016 to 0.057)^c^ | 5.1 |
| Somatic complaints | -0.048 (-0.082 to -0.013)^c^ | -0.003 (-0.009 to 0.004) | 6.3 | 0.724 (0.656 to 0.791)^c^ | 0.024 (0.007 to 0.041)^c^ | 3.3 |
| Social problems | -0.106 (-0.143 to -0.066)^c^ | 0.000 (-0.007 to 0.006) | 0 | 0.705 (0.636 to 0.771)^c^ | 0.038 (0.022 to 0.054)^c^ | 5.4 |
| Thought problems | -0.082 (-0.116 to -0.047)^c^ | -0.002 (-0.009 to 0.004) | 2.4 | 0.717 (0.648 to 0.784)^c^ | 0.028 (0.011 to 0.044)^c^ | 3.9 |
| Attention problems | -0.107 (-0.155 to -0.057)^c^ | -0.004 (-0.013 to 0.005) | 3.7 | 0.716 (0.647 to 0.784)^c^ | 0.015 (0.005 to 0.025)^c^ | 2.1 |
| Rule-breaking behavior | -0.084 (-0.113 to -0.054)^c^ | 0.001 (-0.004 to 0.007) | -1.2 | 0.712 (0.644 to 0.778)^c^ | 0.043 (0.023 to 0.062)^c^ | 6.0 |
| Aggressive behavior | -0.215 (-0.274 to -0.154)^c^ | -0.002 (-0.014 to 0.009) | 0.9 | 0.699 (0.630 to 0.767)^c^ | 0.019 (0.011 to 0.027)^c^ | 2.7 |
| Sluggish cognitive tempo | -0.017 (-0.032 to -0.003)^c^ | 0.002 (-0.001 to 0.006) | -12 | 0.751 (0.685 to 0.816)^c^ | 0.015 (-0.019 to 0.048) | 2.0 |
| Obsessive-compulsive problems | -0.039 (-0.066 to -0.011)^c^ | 0.003 (-0.003 to 0.009) | -7.7 | 0.748 (0.679 to 0.814)^c^ | 0.008 (-0.011 to 0.027) | 1.1 |
| Stress problems | -0.075 (-0.128 to -0.020)^c^ | 0.000 (-0.010 to 0.010) | 0 | 0.703 (0.633 to 0.771)^c^ | 0.021 (0.010 to 0.031)^c^ | 3.0 |
| Depression problems | 0.075 (0.039 to 0.112)^c^ | 0.011 (0.004 to 0.017)^c^ | 15 | 0.726 (0.658 to 0.791)^c^ | 0.028 (0.011 to 0.045)^c^ | 3.9 |
| Anxiety problems | -0.091 (-0.132 to -0.048)^c^ | 0.002 (-0.005 to 0.010) | -2.2 | 0.729 (0.659 to 0.797)^c^ | 0.015 (0.002 to 0.029) | 2.1 |
| Somatic problems | -0.019 (-0.044 to 0.006) | -0.003 (-0.008 to 0.002) | 16 | 0.735 (0.667 to 0.801)^c^ | 0.023 (0.001 to 0.044) | 3.1 |
| ADHD problems | -0.149 (-0.191 to -0.106)^c^ | -0.003 (-0.011 to 0.005) | 2.0 | 0.705 (0.636 to 0.772)^c^ | 0.021 (0.010 to 0.033)^c^ | 3.0 |
| Oppositional defiant problems | -0.099 (-0.130 to -0.068)^c^ | 0.000 (-0.006 to 0.006) | 0 | 0.699 (0.629 to 0.768)^c^ | 0.034 (0.018 to 0.050)^c^ | 4.9 |
| Conduct problems | -0.084 (-0.122 to -0.045)^c^ | -0.001 (-0.007 to 0.006) | 1.2 | 0.721 (0.653 to 0.786)^c^ | 0.033 (0.017 to 0.048)^c^ | 4.6 |

Abbreviations: ADHD, attention-deficit/hyperactivity disorder; BMI, body mass index; CI, confidence interval; RAVLT, Rey Auditory Verbal Learning Test.

^a^ See **Figure** caption in main text for model specification. “Annual change” (left columns) refers to age-related changes (ie, main effect of [age]) at median baseline BMI, median baseline cognition, or zero baseline psychopathology endorsement. “Additional annual change” (right columns) refers to changes associated with per unit increase in baseline BMI, cognition, or psychopathology (ie, [age] × [baseline predictor] interactions) that would add linearly to age-related changes. Highlighted results are the ones associated with significant [age] × [baseline predictor] interactions (see **eTable 1**). These results are illustrated in Johnson-Neyman interaction plots in **eFigures 1** and **2**.

^b^ See notes under **Table** in main text for variable details.

^c^ False discovery rate (FDR)-corrected two-tailed *P* ≤ .05.

**eFigure 1.** Johnson-Neyman interaction plots for significant longitudinal associations between BMI and cognition from late childhood through early adolescence (unstandardized estimates)

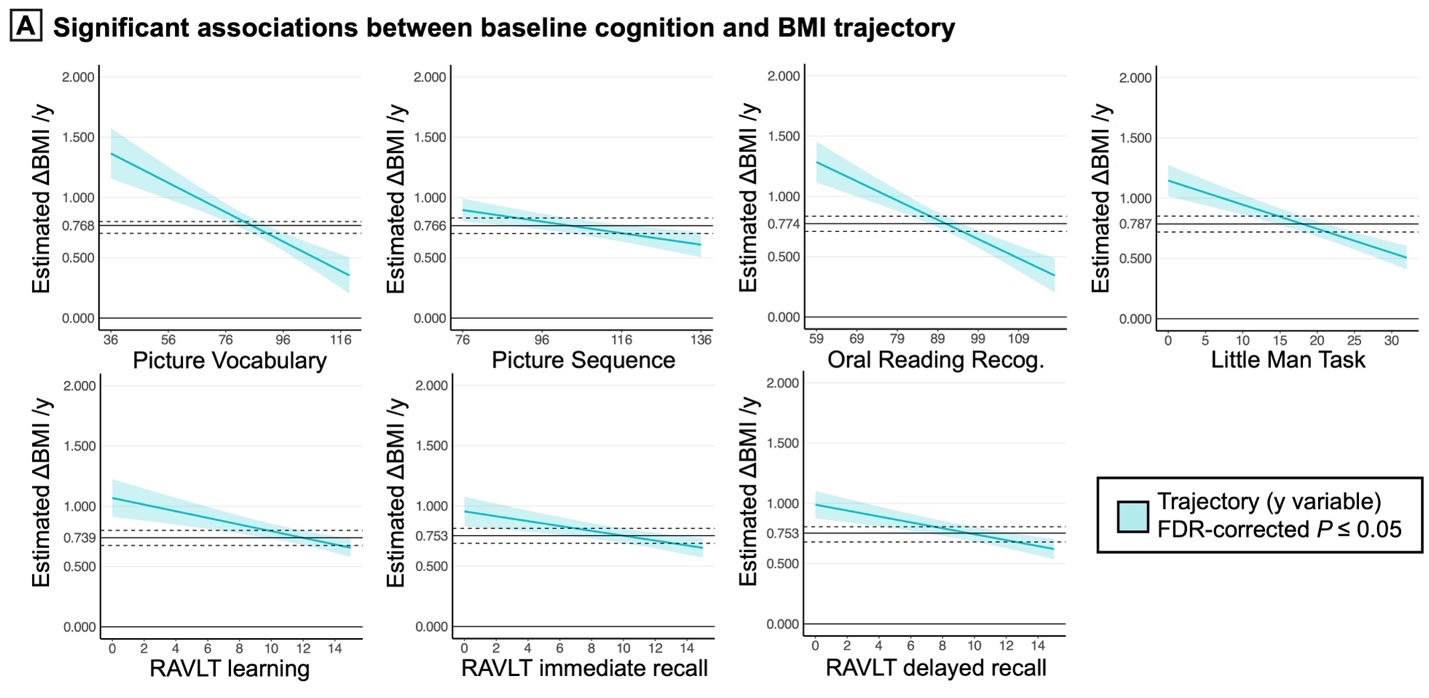

*Caption*. Lower scores on cognitive tests at baseline (x-axis) were associated with greater annual body mass index (BMI) gain (y-axis). Solid horizontal lines represent annual BMI changes corresponding to median baseline cognitive performance, which were proxies of normative changes. Dashed lines and shades represent 95% confidence intervals. These plots visualize the significant unstandardized [age] × [baseline cognition] interactions from **eTable 2**. Here, the [age] × [baseline cognition] interactions were significant for all levels of baseline cognition (blue shades, false discovery rate (FDR)-corrected *P* ≤ .05). RAVLT indicates Rey Auditory Verbal Learning Test.

**eFigure 2.** Johnson-Neyman interaction plots for significant longitudinal associations between BMI and psychopathology from late childhood through early adolescence (unstandardized estimates)

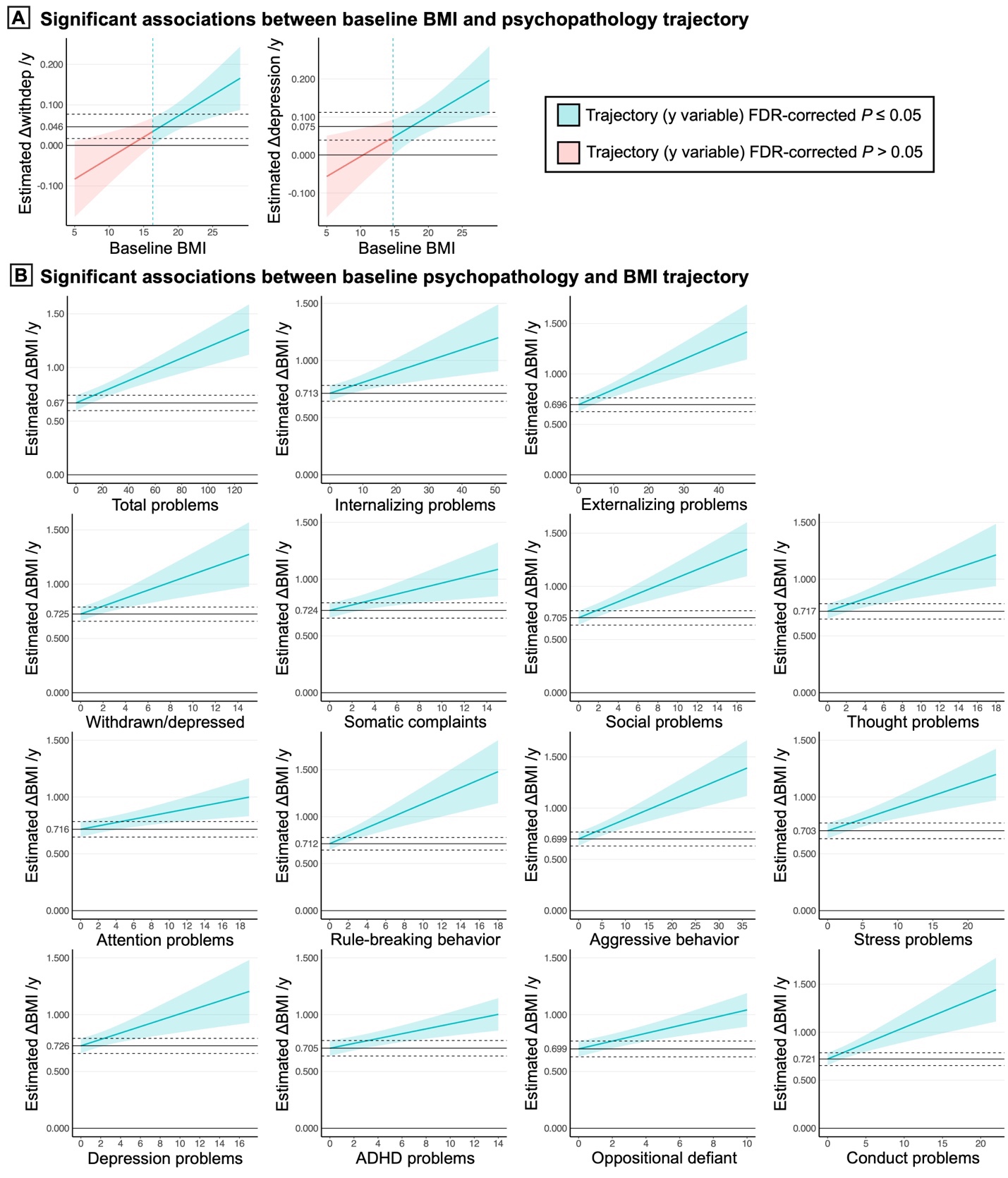

*Caption*. **A**. Higher body mass index (BMI) at baseline (x-axis) was associated with greater increases in withdrawn/depressed (withdep) and depression problems per year (y-axis). Solid horizontal lines represent annual psychopathology changes corresponding to median BMI (17.48 kg/m^2^), which were proxies of normative changes. These plots visualize the significant unstandardized [age] × [baseline BMI] interactions from **eTable 2**. However, the Johnson-Neyman procedure revealed that such interactions were not significant at baseline BMI levels that were slightly below the median (red shades, false discovery rate (FDR)-corrected *P* > .05); ie, psychopathology trajectories only diverged for those with baseline BMI levels near or above the median. **B**. More endorsement of psychopathology at baseline (x-axis) was associated with greater annual BMI gain (y-axis). Solid horizontal lines represent annual BMI changes corresponding to no endorsement at baseline, which were proxies of normative changes. These plots visualize the significant unstandardized [age] × [baseline psychopathology] interactions from **eTable 2**. Here, the [age] × [baseline psychopathology] interactions were significant for all levels of baseline psychopathology endorsement (blue shades, FDR-corrected *P* ≤ .05). In both panels, dashed lines and shades represent 95% confidence intervals. ADHD indicates attention-deficit/hyperactivity disorder.

**eTable 3.** Sex interactions with the longitudinal associations between BMI, cognition, and psychopathology from late childhood through early adolescence^a^

| **Variable^b^** | **Baseline BMI predicting longitudinal cognition or psychopathology**  (ie, [age] × [baseline BMI] interaction) | | | | **Baseline cognition or psychopathology predicting longitudinal BMI**  (ie, [age] × [baseline cognition or psychopathology] interaction) | | | |
| --- | --- | --- | --- | --- | --- | --- | --- | --- |
|  | **Std. *β* (95% CI)** | ***P* value (raw)** | ***P* value (FDR)** | **n** | **Std. *β* (95% CI)** | ***P* value (raw)** | ***P* value (FDR)** | **n** |
| **Cognition** | | | | | | | | |
| Picture Vocabulary | 0.010 (-0.014 to 0.033) | .42 | .74 | 5198 | 0.009 (-0.006 to 0.024) | .24 | .68 | 5212 |
| Flanker Inhibitory Control | -0.003 (-0.034 to 0.029) | .86 | .91 | 5204 | 0.002 (-0.013 to 0.017) | .78 | .88 | 5208 |
| Pattern Comparison | 0.001 (-0.027 to 0.030) | .93 | .93 | 5203 | 0.012 (-0.003 to 0.027) | .11 | .40 | 5201 |
| Picture Sequence | -0.018 (-0.050 to 0.014) | .27 | .68 | 5203 | 0.003 (-0.012 to 0.017) | .73 | .88 | 5209 |
| Oral Reading Recognition | 0.006 (-0.016 to 0.028) | .58 | .87 | 5197 | 0.003 (-0.012 to 0.018) | .66 | .88 | 5204 |
| Little Man Task, n correct | 0.011 (-0.018 to 0.041) | .45 | .74 | 5196 | -0.003 (-0.018 to 0.012) | .72 | .88 | 5082 |
| RAVLT learning | 0.016 (-0.017 to 0.050) | .34 | .69 | 5196 | -0.008 (-0.023 to 0.007) | .31 | .69 | 5193 |
| RAVLT immediate recall | 0.035 (0.002 to 0.068) | .04 | .40 | 5193 | -0.013 (-0.028 to 0.002) | .09 | .40 | 5183 |
| RAVLT delayed recall | 0.027 (-0.004 to 0.059) | .09 | .40 | 5192 | -0.013 (-0.028 to 0.002) | .10 | .40 | 5170 |
| **Psychopathology** | | | | | | | | |
| Total problems | 0.013 (-0.009 to 0.035) | .24 | .70 | 5242 | 0.000 (-0.015 to 0.015) | .99 | .99 | 5269 |
| Internalizing problems | 0.021 (-0.004 to .046) | .09 | .57 | 5242 | -0.005 (-0.020 to 0.010) | .49 | .83 | 5269 |
| Externalizing problems | 0.004 (-0.019 to 0.027) | .74 | .90 | 5242 | 0.010 (-0.006 to 0.025) | .23 | .70 | 5269 |
| Anxious/depressed | 0.028 (0.003 to 0.053) | .03 | .39 | 5242 | -0.004 (-0.019 to 0.010) | .57 | .83 | 5269 |
| Withdrawn/depressed | 0.023 (-0.004 to 0.050) | .10 | .57 | 5242 | -0.001 (-0.017 to 0.014) | .86 | .92 | 5269 |
| Somatic complaints | -0.004 (-0.031 to 0.023) | .76 | .90 | 5242 | -0.005 (-0.020 to 0.010) | .54 | .83 | 5269 |
| Social problems | 0.003 (-0.022 to 0.029) | .78 | .90 | 5242 | -0.001 (-0.017 to 0.014) | .86 | .92 | 5269 |
| Thought problems | 0.007 (-0.018 to 0.032) | .57 | .83 | 5242 | -0.019 (-0.035 to -0.004) | .01 | .39 | 5269 |
| Attention problems | 0.017 (-0.004 to 0.039) | .11 | .57 | 5242 | 0.003 (-0.012 to 0.019) | .66 | .88 | 5269 |
| Rule-breaking behavior | 0.015 (-0.010 to 0.039) | .24 | .70 | 5242 | 0.007 (-0.009 to 0.023) | .40 | .83 | 5269 |
| Aggressive behavior | -0.002 (-0.025 to 0.021) | .87 | .92 | 5242 | 0.010 (-0.005 to 0.025) | .21 | .70 | 5269 |
| Sluggish cognitive tempo | 0.007 (-0.019 to 0.034) | .58 | .83 | 5242 | 0.006 (-0.009 to 0.021) | .42 | .83 | 5269 |
| Obsessive-compulsive problems | 0.026 (0.000 to 0.052) | .05 | .70 | 5242 | -0.004 (-0.019 to 0.010) | .55 | .70 | 5269 |
| Stress problems | 0.012 (-0.012 to 0.036) | .32 | .90 | 5242 | -0.004 (-0.019 to 0.011) | .61 | .78 | 5269 |
| Depression problems | 0.011 (-0.016 to 0.037) | .43 | .39 | 5242 | -0.005 (-0.020 to 0.010) | .48 | .95 | 5269 |
| Anxiety problems | 0.014 (-0.011 to 0.039) | .26 | .89 | 5242 | -0.010 (-0.025 to 0.005) | .20 | .73 | 5269 |
| Somatic problems | -0.004 (-0.032 to 0.024) | .78 | .57 | 5242 | -0.007 (-0.022 to 0.008) | .35 | .63 | 5269 |
| ADHD problems | 0.024 (0.002 to 0.045) | .03 | .83 | 5242 | 0.001 (-0.015 to 0.016) | .92 | .83 | 5269 |
| Oppositional defiant problems | -0.005 (-0.028 to 0.019) | .69 | .51 | 5242 | 0.008 (-0.007 to 0.023) | .29 | .83 | 5269 |
| Conduct problems | 0.020 (-0.005 to 0.044) | .11 | .75 | 5242 | 0.012 (-0.004 to 0.028) | .14 | .84 | 5269 |

Abbreviations: ADHD, attention-deficit/hyperactivity disorder; BMI, body mass index; CI, confidence interval; FDR, false discovery rate; RAVLT, Rey Auditory Verbal Learning Test; Std., standardized.

^a^ Model specification was identical to that provided in **Figure** caption in main text except the estimate of interest was extended from an [age] × [baseline predictor] interaction to an [age] × [baseline predictor] × [sex] interaction.

^b^ See notes under **Table** in main text for variable details.

**eTable 4.** Longitudinal associations between BMI, cognition, and psychopathology in children not using weight-related medications^a^

| **Variable^b^** | **Baseline BMI predicting longitudinal cognition or psychopathology**  (ie, [age] × [baseline BMI] interaction) | | | | **Baseline cognition or psychopathology predicting longitudinal BMI**  (ie, [age] × [baseline cognition or psychopathology] interaction) | | | |
| --- | --- | --- | --- | --- | --- | --- | --- | --- |
|  | **Std. *β* (95% CI)** | ***P* value (raw)** | ***P* value (FDR)** | **n** | **Std. *β* (95% CI)** | ***P* value (raw)** | ***P* value (FDR)** | **n** |
| **Cognition** | | | | | | | | |
| Picture Vocabulary | -0.006 (-0.019 to 0.007) | .36 | .41 | 4516 | -0.023 (-0.031 to -0.015) | < .001 | < .001 | 4530 |
| Flanker Inhibitory Control | -0.013 (-0.030 to 0.004) | .14 | .20 | 4522 | -0.007 (-0.015 to 0.001) | .08 | .14 | 4526 |
| Pattern Comparison | -0.018 (-0.033 to -0.002) | .02 | .05 | 4521 | -0.009 (-0.017 to -0.001) | .02 | .05 | 4523 |
| Picture Sequence | -0.006 (-0.023 to 0.011) | .49 | .52 | 4521 | -0.013 (-0.021 to -0.005) | .001 | .003 | 4527 |
| Oral Reading Recognition | -0.007 (-0.019 to 0.005) | .25 | .32 | 4515 | -0.020 (-0.028 to -0.012) | < .001 | < .001 | 4523 |
| Little Man Task, n correct | 0.009 (-0.006 to 0.025) | .25 | .32 | 4517 | -0.024 (-0.032 to -0.016) | < .001 | < .001 | 4420 |
| RAVLT learning | -0.014 (-0.032 to 0.004) | .14 | .20 | 4515 | -0.017 (-0.025 to -0.009) | < .001 | < .001 | 4516 |
| RAVLT immediate recall | 0.004 (-0.013 to 0.022) | .62 | .62 | 4513 | -0.015 (-0.023 to -0.007) | < .001 | < .001 | 4508 |
| RAVLT delayed recall | 0.009 (-0.008 to 0.026) | .30 | .36 | 4511 | -0.018 (-0.026 to -0.011) | < .001 | < .001 | 4501 |
| **Psychopathology** | | | | | | | | |
| Total problems | 0.000 (-0.013 to 0.012) | .99 | .99 | 4555 | 0.021 (0.013 to 0.029) | < .001 | < .001 | 4580 |
| Internalizing problems | 0.007 (-0.007 to 0.020) | .34 | .55 | 4555 | 0.010 (0.002 to 0.018) | .01 | .03 | 4580 |
| Externalizing problems | -0.002 (-0.015 to 0.010) | .72 | .82 | 4555 | 0.020 (0.012 to 0.028) | < .001 | < .001 | 4580 |
| Anxious/depressed | 0.005 (-0.009 to 0.019) | .49 | .68 | 4555 | 0.005 (-0.003 to 0.012) | .25 | .44 | 4580 |
| Withdrawn/depressed | 0.018 (0.003 to 0.033) | .02 | .05 | 4555 | 0.014 (0.006 to 0.022) | < .001 | .002 | 4580 |
| Somatic complaints | -0.007 (-0.022 to 0.008) | .35 | .55 | 4555 | 0.009 (0.001 to 0.017) | .03 | .07 | 4580 |
| Social problems | -0.004 (-0.018 to 0.010) | .59 | .76 | 4555 | 0.018 (0.010 to 0.026) | < .001 | < .001 | 4580 |
| Thought problems | -0.006 (-0.021 to 0.008) | .37 | .57 | 4555 | 0.011 (0.003 to 0.019) | .005 | .02 | 4580 |
| Attention problems | -0.008 (-0.021 to 0.005) | .21 | .40 | 4555 | 0.012 (0.005 to 0.020) | .002 | .008 | 4580 |
| Rule-breaking behavior | 0.003 (-0.011 to 0.017) | .65 | .78 | 4555 | 0.018 (0.009 to 0.026) | < .001 | < .001 | 4580 |
| Aggressive behavior | -0.005 (-0.018 to 0.008) | .49 | .68 | 4555 | 0.019 (0.011 to 0.027) | < .001 | < .001 | 4580 |
| Sluggish cognitive tempo | 0.005 (-0.009 to 0.019) | .49 | .68 | 4555 | 0.002 (-0.006 to 0.010) | .67 | .78 | 4580 |
| Obsessive-compulsive problems | 0.002 (-0.013 to 0.016) | .79 | .83 | 4555 | 0.001 (-0.007 to 0.009) | .77 | .83 | 4580 |
| Stress problems | -0.004 (-0.018 to 0.010) | .55 | .73 | 4555 | 0.016 (0.008 to 0.024) | < .001 | < .001 | 4580 |
| Depression problems | 0.017 (0.002 to 0.032) | .02 | .06 | 4555 | 0.012 (0.004 to 0.020) | .002 | .008 | 4580 |
| Anxiety problems | 0.004 (-0.010 to 0.018) | .61 | .77 | 4555 | 0.006 (-0.002 to 0.014) | .12 | .27 | 4580 |
| Somatic problems | -0.009 (-0.024 to 0.006) | 0.22 | .40 | 4555 | 0.005 (-0.002 to 0.013) | .18 | .37 | 4580 |
| ADHD problems | -0.008 (-0.021 to 0.005) | .21 | .40 | 4555 | 0.016 (0.008 to 0.024) | < .001 | < .001 | 4580 |
| Oppositional defiant problems | 0.001 (-0.013 to 0.014) | .94 | .96 | 4555 | 0.016 (0.008 to 0.024) | < .001 | < .001 | 4580 |
| Conduct problems | -0.002 (-0.016 to 0.011) | .74 | .82 | 4555 | 0.018 (0.010 to 0.027) | < .001 | < .001 | 4580 |

Abbreviations: ADHD, attention-deficit/hyperactivity disorder; BMI, body mass index; CI, confidence interval; FDR, false discovery rate; RAVLT, Rey Auditory Verbal Learning Test; Std., standardized.

^a^ See **Figure** caption in main text for model specification. Caregivers reported child medication use in the two weeks prior to each study visit. Based on Verhaegen et al.^6^, we screened for use of **antidepressants** (amitriptyline [Elavil^®^], nortriptyline [Pamelor^®^, Aventyl^®^], imipramine [Tofranil^®^], desipramine [Norpramin^®^], doxepin [Sinequan^®^, Silenor^®^], clomipramine [Anafranil^®^], escitalopram [Lexapro^®^], paroxetine [Paxil^®^, Pexeva^®^, Brisdelle^®^], citalopram [Celexa^®^], fluoxetine [Prozac^®^, Rapiflux^®^, Sarafem^®^, Selfemra^®^], sertraline [Zoloft^®^], duloxetine [Cymbalta^®^, Drizalma Sprinkle^®^, Irenka^®^], venlafaxine [Effexor^®^], phenelzine [Nardil^®^], buproprion [Aplenzin^®^, Budeprion^®^, Buproban^®^, Forfivo^®^, Wellbutrin^®^, Zyban^®^], trazodone [Desyrel^®^, Oleptro^®^], nefazodone [Serzone^®^], mirtazapine [Remeron^®^], maprotiline [Ludiomil^®^]), **antipsychotics** (molindone [Moban^®^], haloperidol [Haldol^®^], perphenazine [Etrafon^®^], aripiprazole [Abilify^®^], ziprasidone [Geodon^®^], lurasidone [Latuda^®^], paliperidone [Invega^®^], iloperidone [Fanapt^®^], asenapine [Saphris^®^], amisulpride [Barhemsys^®^], quetiapine [Seroquel^®^], risperidone [Risperdal^®^], clozapine [Clozaril^®^, FazaClo^®^, Versacloz^®^], olanzepine [Zyprexa^®^]), **mood stabilizers** (lithium [Eskalith^®^, Lithobid^®^]), **anticonvulsants** (topiramate [Eprontia^®^, Qudexy^®^, Topamax^®^, Topiragen^®^, Trokendi^®^], zonisamide [Zonegran^®^], lamotrigine [Lamictal^®^], levetiracetam [Elepsia^®^, Keppra^®^], tiagabine [Gabitril^®^], clonazepam [Klonopin^®^], oxcarbazepine [Trileptal^®^], gabapentin [FusePaq Fanatrex^®^, Gabarone^®^, Gralise^®^, Neurontin^®^], pregabalin [Lyrica^®^], valproic acid/ divalproex sodium [Depakene^®^, Depakote^®^, Stavzor^®^], carbamazepine [Carbatrol^®^, Epitol^®^, Equetro^®^, Tegretol^®^]), **ADHD medications** (methylphenidate [Aptensio^®^, Concerta^®^, Cotempla^®^, Jornay^®^, Metadate^®^, Methylin^®^, QuilliChew^®^, Quillivant^®^, Ritalin^®^], dextroamphetamine [Dexedrine^®^, Dextrostat^®^, Liquadd^®^, ProCentra^®^, Zenzedi^®^], dexmethylphenidate [Focalin^®^], amphetamine [Adzenys^®^, Dyanavel^®^, Evekeo^®^], lisdexamfetamine [Vyvanse^®^], combined [Adderall^®^], atomoxetine [Strattera^®^], guanfacine [Intuniv^®^, Tenex^®^]), **growth hormones**, **thyroid hormones** (levothyroxine [Levothroid^®^, Levoxyl^®^, Synthroid^®^, Tirosint^®^, Unithroid^®^]), and **diabetes medications** (insulin, metformin [Fortamet^®^, Glucophage^®^, Glumetza^®^, Riomet^®^]).

^b^ See notes under **Table** in main text for variable details.

**eTable 5.** Longitudinal associations between BMI, cognition, and psychopathology in children without caregiver-reported common psychiatric diagnoses at baseline^a^

| **Variable^b^** | **Baseline BMI predicting longitudinal cognition or psychopathology**  (ie, [age] × [baseline BMI] interaction) | | | | **Baseline cognition or psychopathology predicting longitudinal BMI**  (ie, [age] × [baseline cognition or psychopathology] interaction) | | | |
| --- | --- | --- | --- | --- | --- | --- | --- | --- |
|  | **Std. *β* (95% CI)** | ***P* value (raw)** | ***P* value (FDR)** | **n** | **Std. *β* (95% CI)** | ***P* value (raw)** | ***P* value (FDR)** | **n** |
| **Cognition** | | | | | | | | |
| Picture Vocabulary | -0.005 (-0.018 to 0.008) | .46 | .51 | 4519 | -0.021 (-0.029 to -0.013) | < .001 | < .001 | 4528 |
| Flanker Inhibitory Control | -0.011 (-0.028 to 0.006) | .21 | .28 | 4525 | -0.007 (-0.015 to 0.001) | .09 | .17 | 4524 |
| Pattern Comparison | -0.015 (-0.031 to 0.000) | .05 | .09 | 4524 | -0.009 (-0.016 to -0.001) | .03 | .07 | 4519 |
| Picture Sequence | -0.005 (-0.022 to 0.012) | .55 | .56 | 4524 | -0.012 (-0.020 to -0.004) | .002 | .005 | 4525 |
| Oral Reading Recognition | -0.004 (-0.016 to 0.008) | .51 | .54 | 4518 | -0.019 (-0.027 to -0.011) | < .001 | < .001 | 4520 |
| Little Man Task, n correct | 0.008 (-0.008 to 0.023) | .34 | .41 | 4520 | -0.022 (-0.030 to -0.015) | < .001 | < .001 | 4420 |
| RAVLT learning | -0.012 (-0.030 to 0.006) | .18 | .25 | 4516 | -0.016 (-0.024 to -0.008) | < .001 | < .001 | 4512 |
| RAVLT immediate recall | 0.013 (-0.005 to 0.030) | .15 | .23 | 4515 | -0.015 (-0.022 to -0.007) | < .001 | .001 | 4504 |
| RAVLT delayed recall | 0.013 (-0.004 to 0.029) | .14 | .23 | 4513 | -0.018 (-0.026 to -0.010) | < .001 | < .001 | 4496 |
| **Psychopathology** | | | | | | | | |
| Total problems | 0.002 (-0.010 to 0.015) | .71 | .81 | 4555 | 0.021 (0.013 to 0.029) | < .001 | < .001 | 4578 |
| Internalizing problems | 0.007 (-0.007 to 0.021) | .32 | .54 | 4555 | 0.011 (0.003 to 0.019) | .006 | .02 | 4578 |
| Externalizing problems | 0.000 (-0.013 to 0.013) | .98 | .98 | 4555 | 0.020 (0.012 to 0.028) | < .001 | < .001 | 4578 |
| Anxious/depressed | 0.004 (-0.010 to 0.018) | .53 | .72 | 4555 | 0.005 (-0.003 to 0.013) | .21 | .40 | 4578 |
| Withdrawn/depressed | 0.021 (0.006 to 0.036) | .007 | .02 | 4555 | 0.014 (0.007 to 0.022) | < .001 | .002 | 4578 |
| Somatic complaints | -0.008 (-0.023 to 0.007) | .30 | .52 | 4555 | 0.010 (0.002 to 0.018) | .01 | .03 | 4578 |
| Social problems | -0.005 (-0.019 to 0.009) | .50 | .72 | 4555 | 0.019 (0.011 to 0.027) | < .001 | < .001 | 4578 |
| Thought problems | -0.002 (-0.016 to 0.012) | .76 | .83 | 4555 | 0.010 (0.002 to 0.018) | .01 | .03 | 4578 |
| Attention problems | -0.004 (-0.017 to 0.009) | .54 | .72 | 4555 | 0.013 (0.005 to 0.021) | .002 | .005 | 4578 |
| Rule-breaking behavior | 0.005 (-0.009 to 0.019) | .51 | .72 | 4555 | 0.017 (0.009 to 0.025) | < .001 | < .001 | 4578 |
| Aggressive behavior | -0.003 (-0.016 to 0.010) | .68 | .81 | 4555 | 0.019 (0.011 to 0.027) | < .001 | < .001 | 4578 |
| Sluggish cognitive tempo | 0.009 (-0.006 to 0.023) | .24 | .43 | 4555 | 0.002 (-0.006 to 0.010) | .62 | .79 | 4578 |
| Obsessive-compulsive problems | 0.005 (-0.009 to 0.020) | .47 | .72 | 4555 | 0.001 (-0.006 to 0.009) | .71 | .81 | 4578 |
| Stress problems | -0.002 (-0.016 to 0.012) | .82 | .86 | 4555 | 0.015 (0.007 to 0.023) | < .001 | .001 | 4578 |
| Depression problems | 0.019 (0.004 to 0.034) | .01 | .03 | 4555 | 0.012 (0.004 to 0.019) | .004 | .01 | 4578 |
| Anxiety problems | 0.003 (-0.011 to 0.017) | .63 | .79 | 4555 | 0.005 (-0.002 to 0.013) | .18 | .36 | 4578 |
| Somatic problems | -0.010 (-0.025 to 0.005) | .18 | .36 | 4555 | 0.007 (-0.001 to 0.015) | .09 | .21 | 4578 |
| ADHD problems | -0.004 (-0.017 to 0.009) | .53 | .72 | 4555 | 0.016 (0.008 to 0.024) | < .001 | < .001 | 4578 |
| Oppositional defiant problems | 0.002 (-0.011 to 0.016) | .73 | .81 | 4555 | 0.016 (0.008 to 0.024) | < .001 | < .001 | 4578 |
| Conduct problems | 0.000 (-0.014 to 0.014) | .98 | .98 | 4555 | 0.017 (0.009 to 0.025) | < .001 | < .001 | 4578 |

Abbreviations: ADHD, attention-deficit/hyperactivity disorder; BMI, body mass index; CI, confidence interval; FDR, false discovery rate; RAVLT, Rey Auditory Verbal Learning Test; Std., standardized.

^a^ See **Figure** caption in main text for model specification. Common psychiatric diagnoses included ADHD, depression, bipolar disorder, anxiety, and phobias.

^b^ See notes under **Table** in main text for variable details.

**eTable 6.** Longitudinal associations between WC, cognition, and psychopathology from late childhood through early adolescence (standardized estimates)^a^

| **Variable^b^** | **Baseline WC predicting longitudinal cognition or psychopathology**  (ie, [age] × [baseline WC] interaction) | | | | **Baseline cognition or psychopathology predicting longitudinal WC**  (ie, [age] × [baseline cognition or psychopathology] interaction) | | | |
| --- | --- | --- | --- | --- | --- | --- | --- | --- |
|  | **Std. *β* (95% CI)** | ***P* value (raw)** | ***P* value (FDR)** | **n** | **Std. *β* (95% CI)** | ***P* value (raw)** | ***P* value (FDR)** | **n** |
| **Cognition** | | | | | | | | |
| Picture Vocabulary | -0.006 (-0.018 to 0.006) | .30 | .39 | 5212 | -0.024 (-0.033 to -0.014) | < .001 | < .001 | 5209 |
| Flanker Inhibitory Control | -0.014 (-0.029 to 0.002) | .09 | .16 | 5218 | -0.012 (-0.022 to -0.003) | .009 | .02 | 5205 |
| Pattern Comparison | -0.009 (-0.023 to 0.005) | .22 | .30 | 5217 | -0.011 (-0.021 to -0.002) | .01 | .03 | 5198 |
| Picture Sequence | -0.007 (-0.023 to 0.009) | .37 | .44 | 5217 | -0.016 (-0.025 to -0.007) | < .001 | .003 | 5206 |
| Oral Reading Recognition | -0.005 (-0.016 to 0.006) | .39 | .44 | 5211 | -0.027 (-0.037 to -0.018) | < .001 | < .001 | 5201 |
| Little Man Task, n correct | 0.003 (-0.012 to 0.018) | .69 | .73 | 5210 | -0.017 (-0.027 to -0.008) | < .001 | .001 | 5079 |
| RAVLT learning | -0.003 (-0.020 to 0.014) | .75 | .75 | 5210 | -0.011 (-0.020 to -0.002) | .02 | .04 | 5190 |
| RAVLT immediate recall | 0.011 (-0.005 to 0.028) | .17 | .26 | 5207 | -0.014 (-0.023 to -0.005) | .003 | .01 | 5180 |
| RAVLT delayed recall | 0.013 (-0.002 to 0.029) | .09 | .16 | 5206 | -0.015 (-0.024 to -0.005) | .002 | .007 | 5167 |
| **Psychopathology** | | | | | | | | |
| Total problems | 0.005 (-0.006 to 0.016) | .37 | .62 | 5257 | 0.016 (0.007 to 0.025) | < .001 | .02 | 5266 |
| Internalizing problems | 0.010 (-0.002 to 0.023) | .10 | .28 | 5257 | 0.006 (-0.004 to 0.015) | .24 | .53 | 5266 |
| Externalizing problems | 0.003 (-0.009 to 0.014) | .66 | .74 | 5257 | 0.014 (0.005 to 0.024) | .002 | .02 | 5266 |
| Anxious/depressed | 0.006 (-0.007 to 0.019) | .32 | .62 | 5257 | 0.004 (-0.005 to 0.013) | .41 | .62 | 5266 |
| Withdrawn/depressed | 0.014 (0.000 to 0.028) | .04 | .13 | 5257 | 0.006 (-0.004 to 0.015) | .24 | .53 | 5266 |
| Somatic complaints | 0.003 (-0.010 to 0.017) | .63 | .74 | 5257 | 0.004 (-0.005 to 0.014) | .35 | .62 | 5266 |
| Social problems | -0.001 (-0.014 to 0.012) | .86 | .91 | 5257 | 0.015 (0.006 to 0.024) | .002 | .02 | 5266 |
| Thought problems | -0.005 (-0.018 to 0.007) | .42 | .62 | 5257 | 0.016 (0.007 to 0.025) | < .001 | .02 | 5266 |
| Attention problems | -0.003 (-0.014 to 0.008) | .57 | .69 | 5257 | 0.012 (0.003 to 0.021) | .01 | .04 | 5266 |
| Rule-breaking behavior | 0.007 (-0.005 to 0.020) | .26 | .54 | 5257 | 0.014 (0.004 to 0.023) | .005 | .03 | 5266 |
| Aggressive behavior | -0.001 (-0.012 to 0.011) | .93 | .93 | 5257 | 0.014 (0.004 to 0.023) | .004 | .03 | 5266 |
| Sluggish cognitive tempo | 0.006 (-0.007 to 0.019) | .39 | .62 | 5257 | 0.003 (-0.006 to 0.012) | .52 | .67 | 5266 |
| Obsessive-compulsive problems | 0.004 (-0.009 to 0.017) | .54 | .67 | 5257 | 0.009 (-0.001 to 0.018) | .07 | .19 | 5266 |
| Stress problems | -0.001 (-0.013 to 0.012) | .93 | .93 | 5257 | 0.012 (0.003 to 0.022) | .009 | .04 | 5266 |
| Depression problems | 0.016 (0.002 to 0.029) | .02 | .08 | 5257 | 0.005 (-0.005 to 0.014) | .32 | .62 | 5266 |
| Anxiety problems | 0.005 (-0.008 to 0.017) | .47 | .65 | 5257 | 0.007 (-0.002 to 0.016) | .14 | .35 | 5266 |
| Somatic problems | 0.006 (-0.008 to 0.019) | .43 | .62 | 5257 | 0.004 (-0.005 to 0.013) | .43 | .62 | 5266 |
| ADHD problems | -0.002 (-0.013 to 0.008) | .66 | .74 | 5257 | 0.012 (0.003 to 0.022) | .008 | .04 | 5266 |
| Oppositional defiant problems | 0.004 (-0.008 to 0.016) | .51 | .67 | 5257 | 0.011 (0.001 to 0.020) | .02 | .08 | 5266 |
| Conduct problems | 0.001 (-0.011 to 0.014) | .84 | .91 | 5257 | 0.013 (0.004 to 0.023) | .006 | .03 | 5266 |

Abbreviations: ADHD, attention-deficit/hyperactivity disorder; CI, confidence interval; FDR, false discovery rate; RAVLT, Rey Auditory Verbal Learning Test; Std., standardized; WC, waist circumference.

^a^ See **Figure** caption in main text for model specification.

^b^ See notes under **Table** in main text for variable details.

**eTable 7.** Longitudinal associations between WC, cognition, and psychopathology from late childhood through early adolescence (unstandardized estimates)^a^

| **Variable^b^** | **Baseline WC predicting longitudinal cognition or psychopathology** | | | **Baseline cognition or psychopathology predicting longitudinal WC** | | |
| --- | --- | --- | --- | --- | --- | --- |
|  | **Annual change in cognition or psychopathology at median WC of 25.25 in**  **(95% CI)**  (ie, main effect of [age]) | **Additional annual change in cognition or psychopathology per 1 in increase in WC (95% CI)**  (ie, interaction of [age] × [baseline WC]) | **%** | **Annual change in WC at median cognition or no psychopathology (95% CI)**  (ie, main effect of [age]) | **Additional annual change in WC per 1 point increase in cognition or psychopathology (95% CI)**  (ie, interaction of [age] × [baseline cognition or psychopathology]) | **%** |
| **Cognition** | | | | | | |
| Picture Vocabulary | 2.243 (2.135 to 2.351)^c^ | -0.011 (-0.032 to 0.010) | -0.5 | 0.968 (0.823 to 1.112)^c^ | -0.013 (-0.018 to -0.008)^c^ | -1.3 |
| Flanker Inhibitory Control | 2.743 (2.603 to 2.884)^c^ | -0.025 (-0.053 to 0.004) | -0.9 | 0.954 (0.807 to 1.098)^c^ | -0.006 (-0.011 to -0.002)^c^ | -0.6 |
| Pattern Comparison | 7.048 (6.799 to 7.296)^c^ | -0.031 (-0.081 to 0.019) | -0.4 | 0.955 (0.808 to 1.099)^c^ | -0.003 (-0.006 to -0.001)^c^ | -0.3 |
| Picture Sequence | 3.265 (3.058 to 3.472)^c^ | -0.019 (-0.061 to 0.023) | -0.6 | 0.967 (0.820 to 1.111)^c^ | -0.006 (-0.009 to -0.002)^c^ | -0.6 |
| Oral Reading Recognition | 1.821 (1.738 to 1.904)^c^ | -0.007 (-0.023 to 0.009) | -0.4 | 0.976 (0.832 to 1.118)^c^ | -0.018 (-0.024 to -0.012)^c^ | -1.9 |
| Little Man Task, n correct | 2.284 (2.191 to 2.377)^c^ | 0.004 (-0.015 to 0.023) | 0.2 | 0.993 (0.846 to 1.137)^c^ | -0.014 (-0.021 to -0.006)^c^ | -1.4 |
| RAVLT learning | 0.081 (0.038 to 0.124)^c^ | -0.001 (-0.010 to 0.007) | -1.2 | 0.958 (0.810 to 1.103)^c^ | -0.018 (-0.034 to -0.003)^c^ | -1.9 |
| RAVLT immediate recall | 0.169 (0.122 to 0.218)^c^ | 0.007 (-0.003 to 0.016) | 4.1 | 0.966 (0.820 to 1.111)^c^ | -0.020 (-0.034 to -0.007)^c^ | -2.1 |
| RAVLT delayed recall | 0.082 (0.032 to 0.132)^c^ | 0.009 (-0.001 to 0.019) | 11 | 0.959 (0.812 to 1.103)^c^ | -0.020 (-0.033 to -0.007)^c^ | -2.1 |
| **Psychopathology** | | | | | | |
| Total problems | -0.822 (-1.075 to -0.560)^c^ | 0.020 (-0.024 to 0.063) | -2.4 | 0.894 (0.742 to 1.043)^c^ | 0.004 (0.002 to 0.007)^c^ | 0.4 |
| Internalizing problems | -0.073 (-0.161 to 0.018) | 0.013 (-0.003 to 0.029) | -18 | 0.942 (0.792 to 1.090)^c^ | 0.005 (-0.003 to 0.012) | 0.5 |
| Externalizing problems | -0.296 (-0.378 to -0.213)^c^ | 0.003 (-0.011 to 0.018) | -1.0 | 0.916 (0.767 to 1.063)^c^ | 0.011 (0.004 to 0.019)^c^ | 1.2 |
| Anxious/depressed | -0.077 (-0.125 to -0.027)^c^ | 0.004 (-0.005 to 0.013) | -5.2 | 0.950 (0.801 to 1.097)^c^ | 0.006 (-0.008 to 0.019) | 0.6 |
| Withdrawn/depressed | 0.051 (0.021 to 0.082)^c^ | 0.006 (0.000 to 0.011) | 12 | 0.950 (0.802 to 1.095)^c^ | 0.015 (-0.010 to 0.041) | 1.6 |
| Somatic complaints | -0.049 (-0.083 to -0.015)^c^ | 0.002 (-0.005 to 0.008) | -4.1 | 0.950 (0.801 to 1.096)^c^ | 0.010 (-0.011 to 0.031) | 1.1 |
| Social problems | -0.104 (-0.141 to -0.064)^c^ | -0.001 (-0.007 to 0.006) | 1.0 | 0.921 (0.773 to 1.067)^c^ | 0.031 (0.011 to 0.050)^c^ | 3.4 |
| Thought problems | -0.082 (-0.116 to -0.047)^c^ | -0.003 (-0.009 to 0.004) | 3.7 | 0.911 (0.762 to 1.058)^c^ | 0.035 (0.015 to 0.055)^c^ | 3.8 |
| Attention problems | -0.109 (-0.156 to -0.060)^c^ | -0.002 (-0.011 to 0.006) | 1.8 | 0.919 (0.769 to 1.067)^c^ | 0.016 (0.004 to 0.027)^c^ | 1.7 |
| Rule-breaking behavior | -0.084 (-0.114 to -0.054)^c^ | 0.003 (-0.002 to 0.008) | -3.6 | 0.926 (0.778 to 1.072)^c^ | 0.034 (0.010 to 0.058)^c^ | 3.7 |
| Aggressive behavior | -0.214 (-0.273 to -0.154)^c^ | 0.000 (-0.011 to 0.010) | 0 | 0.920 (0.771 to 1.066)^c^ | 0.014 (0.005 to 0.024)^c^ | 1.5 |
| Sluggish cognitive tempo | -0.016 (-0.030 to -0.001) | 0.001 (-0.002 to 0.004) | -6.2 | 0.957 (0.810 to 1.102)^c^ | 0.013 (-0.028 to 0.055) | 1.4 |
| Obsessive-compulsive problems | -0.038 (-0.065 to -0.009)^c^ | 0.002 (-0.004 to 0.007) | -5.3 | 0.936 (0.788 to 1.083)^c^ | 0.022 (-0.002 to 0.045) | 2.4 |
| Stress problems | -0.074 (-0.126 to -0.020)^c^ | 0.000 (-0.010 to 0.009) | 0 | 0.919 (0.768 to 1.066)^c^ | 0.017 (0.004 to 0.030)^c^ | 1.8 |
| Depression problems | 0.079 (0.043 to 0.116)^c^ | 0.008 (0.001 to 0.014) | 10 | 0.951 (0.803 to 1.097)^c^ | 0.011 (-0.010 to 0.032) | 1.2 |
| Anxiety problems | -0.090 (-0.131 to -0.047)^c^ | 0.003 (-0.005 to 0.010) | -3.3 | 0.939 (0.790 to 1.086)^c^ | 0.013 (-0.004 to 0.030) | 1.4 |
| Somatic problems | -0.022 (-0.047 to 0.002) | 0.002 (-0.003 to 0.007) | -9.1 | 0.953 (0.805 to 1.098)^c^ | 0.011 (-0.016 to 0.037) | 1.2 |
| ADHD problems | -0.150 (-0.191 to -0.108)^c^ | -0.002 (-0.009 to 0.006) | 1.3 | 0.916 (0.766 to 1.064)^c^ | 0.019 (0.005 to 0.033)^c^ | 2.1 |
| Oppositional defiant problems | -0.099 (-0.129 to -0.067)^c^ | 0.002 (-0.004 to 0.007) | -2.0 | 0.924 (0.773 to 1.071)^c^ | 0.023 (0.003 to 0.043)^c^ | 2.5 |
| Conduct problems | -0.085 (-0.123 to -0.045)^c^ | 0.001 (-0.006 to 0.007) | -1.2 | 0.933 (0.786 to 1.078)^c^ | 0.027 (0.008 to 0.046)^c^ | 2.9 |

Abbreviations: ADHD, attention-deficit/hyperactivity disorder; CI, confidence interval; RAVLT, Rey Auditory Verbal Learning Test; WC, waist circumference

^a^ See **Figure** caption in main text for model specification. “Annual change” (left columns) refers to age-related changes (ie, main effect of [age]) at median baseline WC, median baseline cognition, or zero baseline psychopathology endorsement. “Additional annual change” (right columns) refers to changes associated with per unit increase in baseline WC, cognition, or psychopathology (ie, [age] × [baseline predictor] interactions) that would add linearly to age-related changes. Highlighted results are the ones associated with significant [age] × [baseline predictor] interactions (see **eTable 6**).

^b^ See notes under **Table** in main text for variable details.

^c^ False discovery rate (FDR)-corrected two-tailed *P* ≤ .05.

**eTable 8.** Sex interactions with the longitudinal associations between WC, cognition, and psychopathology from late childhood through early adolescence^a^

| **Variable^b^** | **Baseline WC predicting longitudinal cognition or psychopathology**  (ie, [age] × [baseline WC] interaction) | | | | **Baseline cognition or psychopathology predicting longitudinal WC**  (ie, [age] × [baseline cognition or psychopathology] interaction) | | | |
| --- | --- | --- | --- | --- | --- | --- | --- | --- |
|  | **Std. *β* (95% CI)** | ***P* value (raw)** | ***P* value (FDR)** | **n** | **Std. *β* (95% CI)** | ***P* value (raw)** | ***P* value (FDR)** | **n** |
| **Cognition** | | | | | | | | |
| Picture Vocabulary | -0.006 (-0.030 to 0.018) | .62 | .88 | 5212 | -0.001 (-0.019 to 0.017) | .91 | .96 | 5209 |
| Flanker Inhibitory Control | -0.006 (-0.037 to 0.026) | .71 | .88 | 5218 | 0.007 (-0.011 to 0.026) | .44 | .88 | 5205 |
| Pattern Comparison | -0.004 (-0.032 to 0.025) | .79 | .89 | 5217 | 0.008 (-0.010 to 0.026) | .37 | .88 | 5198 |
| Picture Sequence | -0.028 (-0.059 to 0.004) | .09 | .80 | 5217 | -0.003 (-0.021 to 0.015) | .73 | .88 | 5206 |
| Oral Reading Recognition | 0.000 (-0.022 to 0.022) | .98 | .98 | 5211 | 0.009 (-0.009 to 0.028) | .34 | .88 | 5201 |
| Little Man Task, n correct | 0.006 (-0.023 to 0.035) | .71 | .88 | 5210 | -0.003 (-0.022 to 0.015) | .72 | .88 | 5079 |
| RAVLT learning | 0.025 (-0.008 to 0.059) | .14 | .84 | 5210 | 0.005 (-0.013 to 0.023) | .60 | .88 | 5190 |
| RAVLT immediate recall | 0.028 (-0.004 to 0.061) | .09 | .80 | 5207 | -0.006 (-0.025 to 0.012) | .51 | .88 | 5180 |
| RAVLT delayed recall | 0.021 (-0.010 to 0.052) | .19 | .85 | 5206 | -0.005 (-0.023 to 0.014) | .63 | .88 | 5167 |
| **Psychopathology** | | | | | | | | |
| Total problems | 0.015 (-0.007 to 0.037) | .19 | .48 | 5257 | -0.004 (-0.022 to 0.015) | .71 | .86 | 5266 |
| Internalizing problems | 0.019 (-0.006 to 0.043) | .13 | .36 | 5257 | -0.007 (-0.025 to 0.011) | .44 | .75 | 5266 |
| Externalizing problems | 0.001 (-0.021 to 0.024) | .90 | .95 | 5257 | 0.005 (-0.014 to 0.024) | .60 | .83 | 5266 |
| Anxious/depressed | 0.029 (0.004 to 0.053) | .02 | .25 | 5257 | -0.004 (-0.022 to 0.014) | .70 | .86 | 5266 |
| Withdrawn/depressed | 0.018 (-0.009 to 0.045) | .19 | .48 | 5257 | 0.005 (-0.014 to 0.023) | .62 | .83 | 5266 |
| Somatic complaints | -0.005 (-0.032 to 0.021) | .69 | .86 | 5257 | -0.017 (-0.035 to 0.001) | .07 | .25 | 5266 |
| Social problems | -0.001 (-0.026 to 0.024) | .96 | .98 | 5257 | -0.007 (-0.026 to 0.012) | .45 | .75 | 5266 |
| Thought problems | 0.023 (-0.002 to 0.048) | .07 | .25 | 5257 | -0.008 (-0.026 to 0011) | .40 | .73 | 5266 |
| Attention problems | 0.022 (0.001 to 0.043) | .04 | .25 | 5257 | -0.003 (-0.022 to 0.016) | .75 | .86 | 5266 |
| Rule-breaking behavior | 0.023 (-0.002 to 0.048) | .07 | .25 | 5257 | 0.021 (0.001 to 0.040) | .04 | .25 | 5266 |
| Aggressive behavior | -0.008 (-0.031 to 0.015) | .48 | .77 | 5257 | -0.001 (-0.020 to 0.017) | .88 | .95 | 5266 |
| Sluggish cognitive tempo | 0.012 (-0.014 to 0.038) | .38 | .72 | 5257 | -0.006 (-0.024 to 0.012) | .52 | .81 | 5266 |
| Obsessive-compulsive problems | 0.032 (0.006 to 0.059) | .01 | .25 | 5257 | 0.002 (-0.016 to 0.020) | .83 | .93 | 5266 |
| Stress problems | 0.013 (-0.011 to 0.037) | .28 | .61 | 5257 | -0.011 (-0.029 to 0.007) | .24 | .57 | 5266 |
| Depression problems | 0.024 (-0.002 to 0.050) | .07 | .25 | 5257 | -0.005 (-0.023 to 0.014) | .62 | .83 | 5266 |
| Anxiety problems | 0.013 (-0.012 to 0.038) | .32 | .64 | 5257 | -0.010 (-0.028 to 0.008) | .29 | .61 | 5266 |
| Somatic problems | -0.005 (-0.032 to 0.023) | .73 | .86 | 5257 | -0.017 (-0.035 to 0.002) | .08 | .25 | 5266 |
| ADHD problems | 0.030 (0.008 to 0.051) | .006 | .25 | 5257 | 0.000 (-0.018 to 0.019) | .98 | .98 | 5266 |
| Oppositional defiant problems | -0.024 (-0.048 to 0.000) | .05 | .25 | 5257 | -0.005 (-0.023 to 0.013) | .59 | .83 | 5266 |
| Conduct problems | 0.023 (-0.001 to 0.047) | .06 | .25 | 5257 | 0.016 (-0.004 to 0.035) | .12 | .36 | 5266 |

Abbreviations: ADHD, attention-deficit/hyperactivity disorder; CI, confidence interval; FDR, false discovery rate; RAVLT, Rey Auditory Verbal Learning Test; Std., standardized; WC, waist circumference.

^a^ Model specification was identical to that provided in **Figure** caption in main text except the estimate of interest was extended from an [age] × [baseline predictor] interaction to an [age] × [baseline predictor] × [sex] interaction.

^b^ See notes under **Table** in main text for variable details.

**eTable 9.** Longitudinal associations between WC, cognition, and psychopathology in children not using weight-related medications^a^

| **Variable^b^** | **Baseline WC predicting longitudinal cognition or psychopathology**  (ie, [age] × [baseline WC] interaction) | | | | **Baseline cognition or psychopathology predicting longitudinal WC**  (ie, [age] × [baseline cognition or psychopathology] interaction) | | | |
| --- | --- | --- | --- | --- | --- | --- | --- | --- |
|  | **Std. *β* (95% CI)** | ***P* value (raw)** | ***P* value (FDR)** | **n** | **Std. *β* (95% CI)** | ***P* value (raw)** | ***P* value (FDR)** | **n** |
| **Cognition** | | | | | | | | |
| Picture Vocabulary | -0.009 (-0.021 to 0.004) | .18 | .22 | 4529 | -0.025 (-0.035 to -0.015) | < .001 | < .001 | 4528 |
| Flanker Inhibitory Control | -0.021 (-0.038 to -0.004) | .02 | .04 | 4535 | -0.013 (-0.023 to -0.003) | .01 | .03 | 4524 |
| Pattern Comparison | -0.014 (-0.030 to 0.001) | .06 | .11 | 4534 | -0.012 (-0.021 to -0.002) | .02 | .04 | 4521 |
| Picture Sequence | -0.014 (-0.031 to 0.004) | .12 | .16 | 4534 | -0.018 (-0.027 to -0.008) | < .001 | .002 | 4525 |
| Oral Reading Recognition | -0.010 (-0.022 to 0.002) | .12 | .16 | 4528 | -0.024 (-0.034 to -0.014) | < .001 | < .001 | 4521 |
| Little Man Task, n correct | 0.002 (-0.013 to 0.018) | .78 | .78 | 4530 | -0.014 (-0.024 to -0.004) | .006 | .02 | 4418 |
| RAVLT learning | -0.006 (-0.024 to 0.012) | .53 | .57 | 4528 | -0.012 (-0.021 to -0.002) | .02 | .04 | 4514 |
| RAVLT immediate recall | 0.010 (-0.008 to 0.027) | .28 | .32 | 4526 | -0.015 (-0.025 to -0.006) | .002 | .009 | 4506 |
| RAVLT delayed recall | 0.015 (-0.002 to 0.031) | .09 | .13 | 4524 | -0.015 (-0.025 to -0.005) | .003 | .01 | 4499 |
| **Psychopathology** | | | | | | | | |
| Total problems | 0.003 (-0.010 to 0.015) | .69 | .77 | 4569 | 0.018 (0.008 to 0.028) | < .001 | .01 | 4578 |
| Internalizing problems | 0.008 (-0.005 to 0.022) | .22 | .45 | 4569 | 0.007 (-0.003 to 0.017) | .16 | .37 | 4578 |
| Externalizing problems | 0.001 (-0.012 to 0.014) | .87 | .87 | 4569 | 0.014 (0.004 to 0.024) | .006 | .04 | 4578 |
| Anxious/depressed | 0.005 (-0.009 to 0.019) | .51 | .67 | 4569 | 0.005 (-0.005 to 0.014) | .34 | .53 | 4578 |
| Withdrawn/depressed | 0.011 (-0.004 o 0.026) | .14 | .36 | 4569 | 0.008 (-0.002 to 0.018) | .12 | .31 | 4578 |
| Somatic complaints | 0.004 (-0.010 to 0.019) | .56 | .68 | 4569 | 0.005 (-0.005 to 0.015) | .29 | .50 | 4578 |
| Social problems | -0.006 (-0.020 to 0.008) | .42 | .58 | 4569 | 0.013 (0.003 to 0.023) | .01 | .06 | 4578 |
| Thought problems | -0.008 (-0.022 to 0.006) | .26 | .49 | 4569 | 0.019 (0.009 to 0.029) | < .001 | .004 | 4578 |
| Attention problems | -0.004 (-0.017 to 0.009) | .54 | .68 | 4569 | 0.015 (0.005 to 0.025) | .002 | .02 | 4578 |
| Rule-breaking behavior | 0.006 (-0.008 to 0.020) | .36 | .53 | 4569 | 0.011 (0.001 to 0.022) | .03 | .10 | 4578 |
| Aggressive behavior | -0.002 (-0.015 to 0.011) | .80 | .84 | 4569 | 0.014 (0.004 to 0.023) | .007 | .04 | 4578 |
| Sluggish cognitive tempo | 0.007 (-0.007 to 0.021) | .30 | .50 | 4569 | 0.007 (-0.003 to 0.017) | .17 | .37 | 4578 |
| Obsessive-compulsive problems | 0.002 (-0.013 to 0.016) | .80 | .84 | 4569 | 0.012 (0.002 to 0.022) | .02 | .07 | 4578 |
| Stress problems | -0.003 (-0.017 to 0.011) | .67 | .77 | 4569 | 0.015 (0.005 to 0.025) | .003 | .02 | 4578 |
| Depression problems | 0.013 (-0.002 to 0.028) | .08 | .25 | 4569 | 0.007 (-0.003 to 0.016) | .18 | .38 | 4578 |
| Anxiety problems | 0.004 (-0.010 to 0.018) | .59 | .69 | 4569 | 0.008 (-0.001 to 0.018) | .09 | .26 | 4578 |
| Somatic problems | 0.007 (-0.008 to 0.022) | .38 | .55 | 4569 | 0.005 (-0.005 to 0.015) | .30 | .50 | 4578 |
| ADHD problems | -0.006 (-0.019 to 0.007) | .35 | .53 | 4569 | 0.015 (0.005 to 0.025) | .002 | .02 | 4578 |
| Oppositional defiant problems | 0.004 (-0.009 to 0.018) | .52 | .67 | 4569 | 0.010 (0.001 to 0.022) | .04 | .12 | 4578 |
| Conduct problems | -0.001 (-0.019 to 0.012) | .85 | .87 | 4569 | 0.011 (0.001 to 0.022) | .03 | .10 | 4578 |

Abbreviations: ADHD, attention-deficit/hyperactivity disorder; CI, confidence interval; FDR, false discovery rate; RAVLT, Rey Auditory Verbal Learning Test; Std., standardized; WC, waist circumference.

^a^ See **Figure** caption in main text for model specification. Caregivers reported child medication use in the two weeks prior to each study visit. Based on Verhaegen et al.^6^, we screened for use of **antidepressants** (amitriptyline [Elavil^®^], nortriptyline [Pamelor^®^, Aventyl^®^], imipramine [Tofranil^®^], desipramine [Norpramin^®^], doxepin [Sinequan^®^, Silenor^®^], clomipramine [Anafranil^®^], escitalopram [Lexapro^®^], paroxetine [Paxil^®^, Pexeva^®^, Brisdelle^®^], citalopram [Celexa^®^], fluoxetine [Prozac^®^, Rapiflux^®^, Sarafem^®^, Selfemra^®^], sertraline [Zoloft^®^], duloxetine [Cymbalta^®^, Drizalma Sprinkle^®^, Irenka^®^], venlafaxine [Effexor^®^], phenelzine [Nardil^®^], buproprion [Aplenzin^®^, Budeprion^®^, Buproban^®^, Forfivo^®^, Wellbutrin^®^, Zyban^®^], trazodone [Desyrel^®^, Oleptro^®^], nefazodone [Serzone^®^], mirtazapine [Remeron^®^], maprotiline [Ludiomil^®^]), **antipsychotics** (molindone [Moban^®^], haloperidol [Haldol^®^], perphenazine [Etrafon^®^], aripiprazole [Abilify^®^], ziprasidone [Geodon^®^], lurasidone [Latuda^®^], paliperidone [Invega^®^], iloperidone [Fanapt^®^], asenapine [Saphris^®^], amisulpride [Barhemsys^®^], quetiapine [Seroquel^®^], risperidone [Risperdal^®^], clozapine [Clozaril^®^, FazaClo^®^, Versacloz^®^], olanzepine [Zyprexa^®^]), **mood stabilizers** (lithium [Eskalith^®^, Lithobid^®^]), **anticonvulsants** (topiramate [Eprontia^®^, Qudexy^®^, Topamax^®^, Topiragen^®^, Trokendi^®^], zonisamide [Zonegran^®^], lamotrigine [Lamictal^®^], levetiracetam [Elepsia^®^, Keppra^®^], tiagabine [Gabitril^®^], clonazepam [Klonopin^®^], oxcarbazepine [Trileptal^®^], gabapentin [FusePaq Fanatrex^®^, Gabarone^®^, Gralise^®^, Neurontin^®^], pregabalin [Lyrica^®^], valproic acid/ divalproex sodium [Depakene^®^, Depakote^®^, Stavzor^®^], carbamazepine [Carbatrol^®^, Epitol^®^, Equetro^®^, Tegretol^®^]), **ADHD medications** (methylphenidate [Aptensio^®^, Concerta^®^, Cotempla^®^, Jornay^®^, Metadate^®^, Methylin^®^, QuilliChew^®^, Quillivant^®^, Ritalin^®^], dextroamphetamine [Dexedrine^®^, Dextrostat^®^, Liquadd^®^, ProCentra^®^, Zenzedi^®^], dexmethylphenidate [Focalin^®^], amphetamine [Adzenys^®^, Dyanavel^®^, Evekeo^®^], lisdexamfetamine [Vyvanse^®^], combined [Adderall^®^], atomoxetine [Strattera^®^], guanfacine [Intuniv^®^, Tenex^®^]), **growth hormones**, **thyroid hormones** (levothyroxine [Levothroid^®^, Levoxyl^®^, Synthroid^®^, Tirosint^®^, Unithroid^®^]), and **diabetes medications** (insulin, metformin [Fortamet^®^, Glucophage^®^, Glumetza^®^, Riomet^®^]).

^b^ See notes under **Table** in main text for variable details.

**eTable 10.** Longitudinal associations between WC, cognition, and psychopathology in children without caregiver-reported common psychiatric diagnoses at baseline^a^

| **Variable^b^** | **Baseline WC predicting longitudinal cognition or psychopathology**  (ie, [age] × [baseline WC] interaction) | | | | **Baseline cognition or psychopathology predicting longitudinal WC**  (ie, [age] × [baseline cognition or psychopathology] interaction) | | | |
| --- | --- | --- | --- | --- | --- | --- | --- | --- |
|  | **Std. *β* (95% CI)** | ***P* value (raw)** | ***P* value (FDR)** | **n** | **Std. *β* (95% CI)** | ***P* value (raw)** | ***P* value (FDR)** | **n** |
| **Cognition** | | | | | | | | |
| Picture Vocabulary | -0.008 (-0.021 to 0.004) | .19 | .23 | 4530 | -0.024 (-0.034 to -0.014) | < .001 | < .001 | 4526 |
| Flanker Inhibitory Control | -0.021 (-0.038 to -0.004) | .02 | .04 | 4536 | -0.015 (-0.024 to -0.005) | .004 | .01 | 4522 |
| Pattern Comparison | -0.012 (-0.027 to 0.003) | .12 | .16 | 4535 | -0.010 (-0.020 to 0.000) | .04 | .08 | 4517 |
| Picture Sequence | -0.012 (-0.029 to 0.005) | .17 | .21 | 4535 | -0.015 (-0.025 to -0.006) | .002 | .01 | 4523 |
| Oral Reading Recognition | -0.008 (-0.020 to 0.004) | .20 | .23 | 4529 | -0.023 (-0.033 to -0.013) | < .001 | < .001 | 4518 |
| Little Man Task, n correct | 0.002 (-0.014 to 0.017) | .84 | .84 | 4531 | -0.013 (-0.023 to -0.003) | .008 | .02 | 4418 |
| RAVLT learning | -0.005 (-0.023 to 0.014) | .62 | .66 | 4527 | -0.010 (-0.020 to 0.000) | .04 | .08 | 4510 |
| RAVLT immediate recall | 0.016 (-0.002 to 0.033) | .08 | .12 | 4526 | -0.015 (-0.025 to -0.006) | .002 | .01 | 4502 |
| RAVLT delayed recall | 0.017 (0.000 to 0.033) | .05 | .08 | 4524 | -0.014 (-0.024 to -0.004) | .004 | .01 | 4494 |
| **Psychopathology** | | | | | | | | |
| Total problems | 0.004 (-0.009 to 0.017) | .52 | .82 | 4567 | 0.014 (0.004 to 0.024) | .005 | .12 | 4576 |
| Internalizing problems | 0.007 (-0.007 to 0.021) | .31 | .65 | 4567 | 0.004 (-0.005 to 0.014) | .38 | .68 | 4576 |
| Externalizing problems | 0.003 (-0.010 to 0.015) | .70 | .82 | 4567 | 0.012 (0.002 to 0.022) | .01 | .12 | 4576 |
| Anxious/depressed | 0.002 (-0.012 to 0.015) | .82 | .87 | 4567 | 0.002 (-0.008 to 0.012) | .67 | .82 | 4576 |
| Withdrawn/depressed | 0.013 (-0.011 to 0.018) | .09 | .28 | 4567 | 0.008 (-0.002 to 0.018) | .13 | .34 | 4576 |
| Somatic complaints | 0.003 (-0.011 to 0.018) | .67 | .82 | 4567 | 0.002 (-0.007 to 0.012) | .64 | .82 | 4576 |
| Social problems | -0.008 (-0.022 to 0.006) | .26 | .57 | 4567 | 0.013 (0.003 to 0.023) | .01 | .12 | 4576 |
| Thought problems | -0.004 (-0.018 to 0.010) | .59 | .82 | 4567 | 0.012 (0.002 to 0.022) | .02 | .12 | 4576 |
| Attention problems | 0.002 (-0.011 to 0.015) | .79 | .87 | 4567 | 0.011 (0.002 to 0.021) | .02 | .12 | 4576 |
| Rule-breaking behavior | 0.010 (-0.004 to 0.024) | .17 | .41 | 4567 | 0.010 (0.001 to 0.020) | .04 | .18 | 4576 |
| Aggressive behavior | -0.001 (-0.014 to 0.012) | .85 | .87 | 4567 | 0.012 (0.002 to 0.022) | .02 | .12 | 4576 |
| Sluggish cognitive tempo | 0.010 (-0.004 to 0.025) | .15 | .39 | 4567 | 0.003 (-0.007 to 0.013) | .55 | .82 | 4576 |
| Obsessive-compulsive problems | 0.003 (-0.011 to 0.017) | .67 | .82 | 4567 | 0.008 (-0.002 to 0.018) | .11 | .31 | 4576 |
| Stress problems | -0.002 (-0.016 to 0.012) | .79 | .87 | 4567 | 0.011 (0.002 to 0.021) | .02 | .12 | 4576 |
| Depression problems | 0.013 (-0.002 to 0.028) | .08 | .28 | 4567 | 0.004 (-0.006 to 0.013) | .48 | .80 | 4576 |
| Anxiety problems | 0.003 (-0.011 to 0.0178) | .66 | .82 | 4567 | 0.004 (-0.005 to 0.014) | .38 | .68 | 4576 |
| Somatic problems | 0.006 (-0.009 to 0.020) | .47 | .80 | 4567 | 0.003 (-0.007 to 0.012) | .58 | .82 | 4576 |
| ADHD problems | 0.001 (-0.012 to 0.014) | .86 | .87 | 4567 | 0.012 (0.002 to 0.022) | .02 | .12 | 4576 |
| Oppositional defiant problems | 0.007 (-0.007 to 0.020) | .33 | .66 | 4567 | 0.009 (-0.001 to 0.018) | .08 | .28 | 4576 |
| Conduct problems | 0.001 (-0.013 to 0.015) | .87 | .87 | 4567 | 0.009 (-0.001 to 0.019) | .07 | .26 | 4576 |

Abbreviations: ADHD, attention-deficit/hyperactivity disorder; CI, confidence interval; FDR, false discovery rate; RAVLT, Rey Auditory Verbal Learning Test; Std., standardized; WC, waist circumference.

^a^ See **Figure** caption in main text for model specification. Common psychiatric diagnoses included ADHD, depression, bipolar disorder, anxiety, and phobias.

^b^ See notes under **Table** in main text for variable details.
